# Profiling the genome and proteome of metabolic dysfunction-associated steatotic liver disease identifies potential therapeutic targets

**DOI:** 10.1101/2023.11.30.23299247

**Authors:** Jun Liu, Sile Hu, Lingyan Chen, Charlotte Daly, Cesar Augusto Prada Medina, Tom G Richardson, Matthew Traylor, Niall J Dempster, Richard Mbasu, Thomas Monfeuga, Marijana Vujkovic, Philip S Tsao, Julie A Lynch, Benjamin F. Voight, Kyong-Mi Chang, VA Million Veteran Program, Jeremy F Cobbold, Jeremy W Tomlinson, Cornelia M van Duijn, Joanna M M Howson

## Abstract

**BACKGROUND & AIMS:** Metabolic dysfunction-associated steatotic liver disease (MASLD) affects over 25% of the population and currently has no effective treatments. Plasma proteins with causal evidence may represent promising drug targets. We aimed to identify plasma proteins in the causal pathway of MASLD and explore their interaction with obesity.

**METHODS:** We analysed 2,941 plasma proteins in 43,978 European participants from UK Biobank. We performed genome-wide association study (GWAS) for all MASLD-associated proteins and created the largest MASLD GWAS (109,885 cases/1,014,923 controls). We performed Mendelian Randomization (MR) and integrated proteins and their encoding genes in MASLD ranges to identify candidate causal proteins. We then validated them through independent replication, exome sequencing, liver imaging, bulk and single-cell gene expression, liver biopsies, pathway, and phenome-wide data. We explored the role of obesity by MR and multivariable MR across proteins, body mass index, and MASLD.

**RESULTS:** We found 929 proteins associated with MASLD, reported five novel genetic loci associated with MASLD, and identified 17 candidate MASLD protein targets. We identified four novel targets for MASLD (CD33, GRHPR, HMOX2, and SCG3), provided protein evidence supporting roles of AHCY, FCGR2B, ORM1, and RBKS in MASLD, and validated nine previously known targets. We found that CD33, FCGR2B, ORM1, RBKS, and SCG3 mediated the association of obesity and MASLD, and HMOX2, ORM1, and RBKS had effect on MASLD independent of obesity.

**CONCLUSIONS:** This study identified new protein targets in the causal pathway of MASLD, providing new insights into the multi-omics architecture and pathophysiology of MASLD. These findings advise further therapeutic interventions for MASLD.

## Introduction

Metabolic dysfunction-associated steatotic liver disease (MASLD), formerly known as non-alcoholic fatty liver disease (NAFLD)^1^, is characterised by an excessive accumulation of fat in the liver (>5%) that is not caused by excessive alcohol consumption or other known liver disease aetiologies. Its estimated to affect more than 25% of the global population, making it one of the most common liver diseases worldwide^2^. Its prevalence is increasing rapidly, which is attributed to the increase in obesity. MASLD is a major risk factor for liver fibrosis, metabolic dysfunction associated steatohepatitis (MASH), formerly known as non-alcoholic steatohepatitis (NASH), cirrhosis, liver failure, and liver cancer in Western countries^2^. Currently, there are no specific medications for MASLD, and lifestyle modifications are the mainstay of treatment, which is not achievable by many patients. Thus, there is an urgent need to improve our understanding of MASLD and identify and evaluate therapeutic targets or interventions to treat it more effectively.

Over 95% of all currently known drugs target proteins^3^, highlighting the importance of understanding the role of proteins in the development of MASLD. Understanding how proteins relate to MASLD, especially distinguishing the proteins in the causal pathway of MASLD, could provide insights into the underlying mechanisms of the disease and identify potential therapeutic targets for drug development. Studying liver tissue for proteomic analysis in MASLD is challenging due to a paucity of liver tissue samples available. The liver is the central organ producing and metabolising plasma proteins. Circulating plasma proteins can be an informative read-out of liver function for the discovery phase, followed by a validation in tissue specific biological data such as liver imaging, transcriptomics and biopsy.

The present study aims to identify proteins in the causal pathway of MASLD by analyzing data on 2,941 plasma proteins measured by Olink® in 43,978 European participants in UK Biobank. We find that 929 proteins (31.6%) are observationally associated with MASLD and identify 17 proteins that may be on the causal pathway to MASLD based on Mendelian randomization (MR) and genetics data. These include CD33, GRHPR, HMOX2, and SCG3 which are reported as protein/genetic targets of MASLD or NAFLD for the first time, and AHCY, FCGR2B, ORM1, and RBKS which are identified on their protein levels with MASLD or NAFLD for the first time. We further confirmed their validity using a range of independent methods, including exome sequencing, liver imaging, bulk and single-cell gene expression, liver biopsies, pathway analysis, and phenome-wide data.

## Materials and Methods

### Study participants

We conducted our study within UK Biobank which comprised over 500,000 participants aged 37 to 73 years during the recruitment period (2006 to 2010). Participant data include genome-wide genotyping, exome sequencing, whole-body magnetic resonance imaging, electronic health record linkage, blood and urine biomarkers, and physical and anthropometric measurements. Further details are available online^4^. All participants provided electronically signed informed consent. UK Biobank has approval from the North West Multi-centre Research Ethics Committee, the Patient Information Advisory Group, and the Community Health Index Advisory Group. The current study is a part of UK Biobank project 53639 and 65851.

Human liver biopsies were obtained from female patients undergoing elective surgery, including bariatric procedures and gall bladder removal. All participants gave full, informed, written consent for the liver biopsy (NHS research ethics reference: 17/WM/0130). Clinical and biochemical data were collected on the day of surgery; 36 female European participants were included in the analysis (Supplementary Table 2). An experienced histopathologist assessed all liver biopsies for steatosis and MASLD activity score according to the Kleiner classification^5^.

### Phenotype definitions

We defined MASLD cases based on magnetic resonance imaging-estimated proton density fat fraction (MRI-PDFF)^6^ ≥5% (∼42K participants available) or from primary care records (∼250K participants available), hospital admission or death registration (∼500K participants available) using International Classification of Diseases (ICD)9 (code 5715 and 5718) and ICD10 (code K758, K760 and K746)^7^ until November 2022. We excluded participants with excessive alcohol consumption or any secondary causes of hepatic steatosis (Supplementary Table 3)^8^ from both cases and controls of MASLD analyses. The definition of other traits and covariates are descripted in Supplementary Table 4.

### Genomic data processing

UK Biobank array genotyping was conducted using bespoke Affymetrix UK BiLEVE Axiom® Array or UK Biobank Axiom® array. All genetic data were quality controlled and imputed as described previously^9^. Participants who had gender mismatch, failed quality control, significant missing data, or had heterozygosity were excluded following UK Biobank’s recommendation^10^. Non-European participants were excluded from current analysis. Whole-exome sequencing was measured in 454,787 participants from UK Biobank using a previously described method^11^.

### Proteome measurement

High-throughput proteomics measures were performed by Olink® in a randomly selected 46,673 participants, of which 43,978 Europeans were included in the current proteomics analyses. Details of the Olink proteomics assay, data processing and quality control were descripted previously^12^. We included 2,941 protein variables, of which 12 molecules mapped to two or more possible proteins were analysed as one variable, and 18 proteins duplicated among panels were analysed separately in our analysis. A general map of the proteomics is shown in Supplementary Table 1. The protein values were rank-based inverse normal transformed to ensure that the results were comparable across the entire dataset. Analysis of proteins in the liver biopsies was performed by liquid chromatography-tandem mass spectrometry (LC-MS/MS), which is fully described in Supplemental Materials.

### Statistical analysis

All analyses were performed in R statistical software (version 4.0.3) unless otherwise specified. Two-tailed tests were considered. Detailed data preparation and missing imputation is in Supplemental Materials.

#### Proteome associations

For the association of proteins and outcomes, we used logistic regression for binary outcomes and linear regression for continuous outcomes for each protein separately. We used two models for the regression analysis in UK Biobank: model 1 included age, age squared, sex, fasting time, and batches of proteomic measurement; model 2 included the covariates in model 1 and common lifestyle factors, including smoking status, number of pack-years of smoking, grams of alcohol consumption per week, education, and physical activity. Multiple testing was considered by Matrix Spectral Decomposition (MSD) method^13^. The explanation of each significance threshold in our study is descripted in Supplemental Materials. Linear regression analysis was used to examine the association between specific proteins and features of hepatic steatosis in liver biopsy tissue. The covariates adjusted for in the model included age, and study groups.

We further explored the full STRING protein-protein networks and enrichment facilities of the 929 proteins for MASLD through STRING database^14^ using the high confidence (≥ 0.90) based on experiments or databases source (false discovery rate, *FDR*<0.05). The network was clustered into ten clusters based on KMeans clustering. STRING database included 905 proteins available for MASLD.

#### Genome-wide association study (GWAS) and meta-analysis

We performed GWAS for all proteins (n=41,286), PDFF (n=38,174), and MASLD (11,947 cases and 313,042 controls) in UK Biobank using a whole-genome regression approach implemented in REGENIE^15^. It considers relatedness, population structure, polygenicity, and unbalanced binary traits by Firth logistic regression. We used participants with European ancestry based on both questionnaires and their genetic background. The relatedness included in the analysis was adjusted for using the genotype relatedness matrix. Covariates in the model included age, age squared, sex, the interaction of age and sex, the interaction of age squared and sex, batch and chip of genotyping process, and the first ten principal components. For MASLD GWAS sources, we also included three previous studies: (1) GWAS_Ghodsian_, conducted by N Ghodsian, et al^16^; (2) GWAS_Finngen_, conducted using Finngen R9^17^; and (3) GWAS_MVP_, conducted by M Vujkovic, et al, using unexplained chronic alanine aminotransferase (ALT) elevation as a proxy for MASLD^18^. By METAL^19^, considering study specific weight, genomic control, and sample overlap, we incorporated a meta-analysis of GWAS_Ghodsian_, GWAS_Finngen_, and GWAS_UKB_, correcting for 16.3% overlapped cases and 64.4% overlapped controls(GWAS_meta_) and a meta-analysis of GWAS_Ghodsian_, GWAS_Finngen_, GWAS_UKB_, and GWAS_MVP_, correcting for 2.9% overlapped cases and 56.3% overlapped controls (GWAS_meta+MVP_). GWAS summary statistics for Somalogic proteome were obtained from previous publication which included 35,559 Icelanders^20^. GWAS summary statistics for liver volume, liver iron content, liver fat percentage, and visceral adipose tissue volume were obtained from previous publication which considered over 38,000 participants in UK Biobank^21^.

#### MR, multivariable MR, phenome-wide MR (PheMR) and colocalization

We selected cis-protein quantitative trait loci (pQTLs) (*p*<5×10^−8^, minor allele frequency>1.0×10^−3^), cis-expression quantitative trait loci (eQTLs) (*p*<5×10^−8^, minor allele frequency>0.01) or genetic determinants of BMI (*p*<5×10^−8^, minor allele frequency>0.01) from the GWAS summary statistics^22,23^ based on +/-500kbs of the encoding gene and r^2^<0.01 using a reference panel with 10,000 random European individuals from UK Biobank^24^. We used R package *TwoSampleMR, MendelianRandomization,* and *MRPRESSO* to perform different MR methods, including inverse-variance weighted, weighted mode, weighted median, simple mode, and MR-Egger, Cochran’s Q statistic for heterogeneity effect, MR-Egger intercept test and regression^25^, and MR-PRESSO^26^ for pleiotropic effect, and contamination mixture model^27^ for valid instrumental variables, when allowed.

Multivariable MR was performed using R package *GRAPPLE* with the function *grappleRobustEst* to estimate the causal effects of individual proteins and body mass index (BMI) under a random effect model of the pleiotropic effects^28^. For PheMR, we used the most completed GWAS summary statistics from Pan-UKB team^29^ for 916 selected diseases, which include European cases based on three-digit ICD10 codes. Colocalization analysis was based on *HyPrColoc*^30^. We used non-uniform priors, including genetic variants +/− 500kbs of the encoding genes, and investigated posterior alignment probabilities. We assumed a probability of ≥ 0.7 to indicate significant colocalization.

#### Whole exome sequencing analyses

Exome-wide association analysis was performed through REGENIE^15^ in the European population only to exclude the ancestry heterogeneity as previous publication^11^. Variants were annotated using Variant Effect Predictor (VEP) versions, and Loss-Of-Function Transcript Effect Estimator (LOFTEE) plug-ins, to select the most severe consequence of each variant among all protein-coding transcripts. We focused on variants that are highly or moderately like to influence the function of phenotypes defined through snpEff^31^, including transcript ablation/amplification, splice acceptor/donor variants, loss/gain-of-function mutations, missense variants, inframe insertion or deletion, and protein-altering variants.

#### Gene expression related analyses

Default FUMA^32^ pipeline based on GTEx (v8) dataset^22^ were used to demonstrate the expression levels of the encoding genes of target proteins. It included the gene expression levels through log2 transformed average expression values and enrichment analysis based on pre-calculated differentially expressed genes (DEG) sets across 54 different general tissue types. Genes with log2 transformed expression values greater than 2.84, which corresponds to an expression level approximately 6.5-fold higher than the median expression level across all genes, were considered to be highly expressed. The GWAS summary statistics of gene expression in the liver and adiposity tissues were obtained from GTEx (v8) dataset^22^ and STARNET dataset^33^ and used to identify the genetic variants/determinants of the gene expression (*p*<5×10^−8^).

The bulk RNA-seq data of liver biopsies were obtained from a previous transcriptomic study, which have snap-frozen biopsies from 206 MASLD patients and 10 healthy obese control cases without any biochemical or histological evidence of MASLD processed for RNA sequencing on the Illumina NextSeq 500 system^34^. Sixteen of the 17 candidate causal proteins were available, except for SCG3 which was not measured in the full set of control groups (n<10). *GEO2R* platform was used to perform the analysis^35^.

#### Analysis of *single nucleus RNA sequence* (snRNA-seq) and *single cell RNA sequence* (scRNA-seq)

We integrated six independent studies of liver cell snRNA-seq and scRNA-seq data, considering the differences in sample processing techniques and donor health conditions across the studies^36–42^. Detailed procedure is descripted in Supplemental Materials. For each the liver comprising cell types, we calculated the fraction of cells and the mean expression levels of the encoding genes of target proteins. For genes detected in more than 15% of the cells and a mean expression level greater than 75% of the genes, we compared the differential gene expression levels through the Mann-Whitney-Wilcoxon test across disease groups. We used data from Ramachandran et al.’s study^38^ for MASLD and non-MASLD groups. As there were no available MASLD data for hepatocytes and stellate cells, we used alternative data from NASH or alcoholic liver disease studies^38–40^.

#### Pathway enrichment analyses

Comprehensive enrichment analyses among all the potential causal targets of MASLD were performed through Metascape pipeline^43^. Functional enrichment analyses have been carried out by hypergeometric test and Benjamini-Hochberg p-value correction algorithm with the following ontology sources: GO Biological Processes, DisGeNET^44^, and PaGenBase^45^. All genes in the genome have been used as the enrichment background. Terms with a *p* < 0.01, a minimum count of three, and an enrichment factor > 1.5 (the enrichment factor is the ratio between the observed counts and the counts expected by chance) were collected and grouped into clusters based on their membership similarities.

## Results

A flow chart of the study design is presented in Figure 1.

**Figure 1.**
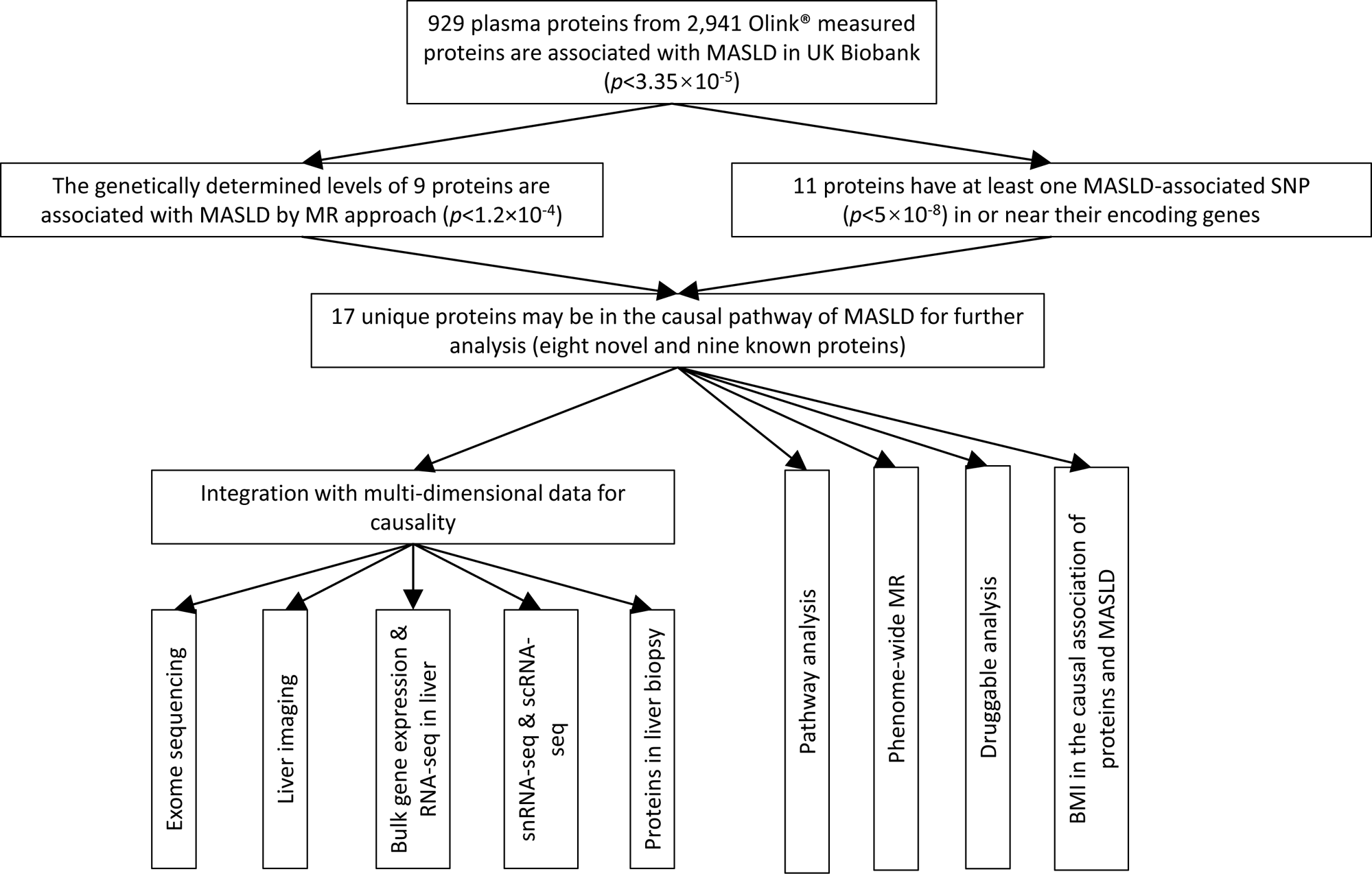
Flow chart of the study design.

### Cohort description

We studied 43,978 randomly selected European participants from UK Biobank for proteomic analyses. After excluding participants with excessive alcohol consumption or secondary liver disease, 1,181 MASLD patients and 30,719 controls were identified. Individuals with MASLD had a higher prevalence of obesity, male gender, diabetes, hypertension, and various metabolic abnormalities compared to controls (*p*<1.0×10^−3^; Table 1). Only 8.4% of participants (3,676 out of 43,978) had liver imaging data, while a comparison between those with and without liver imaging data shows a high correlation in the association of MASLD with its common risk factors (r=0.89, *p*=2.4×10^−11^; Supplementary Figure 1; Supplementary Table 5), suggesting no evidence of major selection bias in the patients included.

**Table 1.** Characteristics of the study population for proteomic analyses.

|  | Total (n = 30,719) | Controls (n = 29,538) | Cases (n = 1,181) |
| --- | --- | --- | --- |
| Age (years), Mean ± SD | 56.98 ± 8.07 | 57 ± 8.09 | 56.63 ± 7.46 |
| Male, n (%) | 12,534 (40.82) | 11,943 (40.43) | 591 (50.04) |
| BMI (kg/m<sup>2</sup>), Mean ± SD | 27.35 ± 4.84 | 27.23 ± 4.79 | 30.30 ± 5.15 |
| Waist circumference (cm), Mean ± SD | 89.49 ± 13.52 | 89.15 ± 13.42 | 98.04 ± 13.20 |
| Smoking status, n (%) |  |  |  |
| Never | 18,354 (59.75) | 17,681 (59.86) | 673 (56.99) |
| Previous | 9,796 (31.89) | 9,385 (31.77) | 411 (34.80) |
| Current | 2,569 (8.36) | 2,472 (8.37) | 97 (8.21) |
| Smoking pack years<sup>a</sup>, Median (IQR) | 9.0 (0.025, 23.4) | 8.75 (0.025, 23.25) | 11.25 (0.025, 26.06) |
| Alcohol grams per week<sup>b</sup>, Median (IQR) | 80.0 (48.0, 118.4) | 80.0 (48.0, 118.40) | 84.0 (56.20, 124.0) |
| Education, n (%) |  |  |  |
| College or University degree | 9,847 (32.06) | 9,492 (32.13) | 355 (30.06) |
| A levels AS levels or equivalent | 3,507 (11.42) | 3,361 (11.38) | 146 (12.36) |
| CSEs or equivalent | 1,661 (5.41) | 1,597 (5.41) | 64 (5.42) |
| NVQ or HND or HNC or equivalent | 1,956 (6.37) | 1,875 (6.35) | 81 (6.86) |
| O levels GCSEs or equivalent | 6,623 (21.56) | 6,367 (21.56) | 256 (21.68) |
| Other professional qualifications | 1,637 (5.33) | 1,563 (5.29) | 74 (6.27) |
| None | 5,488 (17.87) | 5,283 (17.89) | 205 (17.36) |
| Physical activity, n (%) |  |  |  |
| High | 12,328 (40.13) | 11,939 (40.42) | 389 (32.94) |
| Moderate | 12,576 (40.94) | 12,086 (40.92) | 490 (41.49) |
| Low | 5,815 (18.93) | 5,513 (18.66) | 302 (25.57) |
| Glucose (mmol/L), Mean ± SD | 5.07 ± 0.92 | 5.06 ± 0.91 | 5.29 ± 1.22 |
| HbA1c (mmol/mol), Mean ± SD | 36.01 ± 5.66 | 35.93 ± 5.55 | 37.90 ± 7.56 |
| Type 2 diabetes, n (%) | 1,589 (5.17) | 1,437 (4.86) | 152 (12.87) |
| Anti-diabetic medications, n (%) | 1,086 (3.54) | 988 (3.34) | 98 (8.3) |
| Hypertension, n (%) | 15,799 (53.8) | 15,085 (53.42) | 714 (63.19) |
| Systolic blood pressure (mmHg), Mean ± SD | 136.98 ± 18.51 | 136.91 ± 18.55 | 138.66 ± 17.45 |
| Diastolic blood pressure (mmHg), Mean ± SD | 81.56 ± 10.08 | 81.48 ± 10.07 | 83.7 ± 9.89 |
| Anti-hypertensive medications, n (%) | 6,949 (22.62) | 6,570 (22.24) | 379 (32.09) |
| Triglycerides (mmol/L), Median (IQR) | 1.48 (1.05, 2.12) | 1.46 (1.04, 2.1) | 1.9 (1.41, 2.73) |
| HDL-cholesterol (mmol/L), Mean ± SD | 1.43 ± 0.37 | 1.44 ± 0.37 | 1.25 ± 0.31 |
| Lipid-lowering medications, n (%) | 6,308 (20.53) | 5,991 (20.28) | 317 (26.84) |
| AST (U/L), Median (IQR) | 24.1 (20.8, 28.3) | 24 (20.8, 28.1) | 26.3 (22.2, 32.4) |
| ALT (U/L), Median (IQR) | 19.56 (15.12, 26.29) | 19.38 (15.01, 25.92) | 25.97 (19.18, 37.27) |
| GGT (U/L), Median (IQR) | 24.3 (17.6, 36.4) | 24 (17.5, 35.9) | 33.6 (23.8, 52.9) |
| CRP (mg/L), Median (IQR) | 1.32 (0.65, 2.75) | 1.29 (0.64, 2.71) | 1.93 (1.04, 3.83) |
| MRI-PDFF (%), Median (IQR) | 2.9 (2.1, 5) | 2.49 (1.99, 3.16) | 8.6 (6.1, 13.1) |

<sup>a</sup>: Never smokers were not considered when calculating pack years. <sup>b</sup>: Never drinkers were not considered when calculating alcohol grams per week. SD: standard deviation; BMI: body mass index; IQR: interquartile range; AS: Advanced Subsidiary; CES: Certificate of Secondary Education; NVQ: National Vocational Qualification; HND: Higher National Diploma; HNC: Higher National Certificates; GCSE: General Certificate of Secondary Education; HbA1c: Haemoglobin A1c; HDL: high-density lipoprotein; AST: Aspartate aminotransferase; ALT: Alanine aminotransferase; GGT: $\gamma$ -glutamyl transferase; CRP: c-reactive protein; MRI-PDFF: Proton Density Fat Fraction measured by magnetic resonance imaging (MRI).

### Proteins associated with MASLD

After adjusted significance threshold for multiple testing (*p*<3.35×10^−5^), 1,022 (34.8%) of the 2,941 plasma proteins were found to be associated with MASLD based on the baseline model (Supplementary Table 6). Adjustment for common lifestyle factors additionally reduced the number of proteins associated with MASLD by 9.1% to 929 (Figure 2A, Supplementary Figure 2). BMI, the most important risk factor for MASLD, was found to be significantly associated with most of the proteins associated with MASLD (99.0%) apart from nine proteins (Supplementary Table 6; Figure 2B).

**Figure 2.**
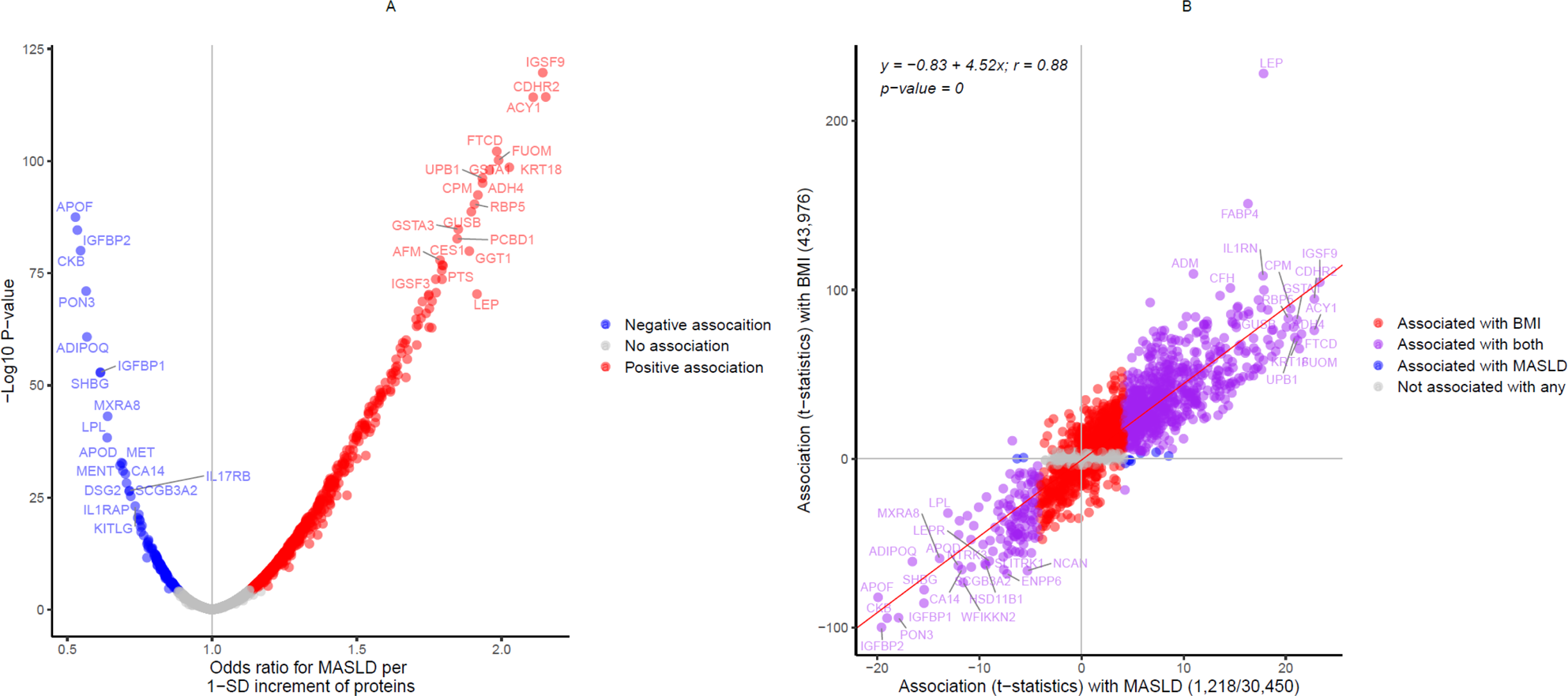
Association of proteins with MASLD and BMI. **A)** Proteome-wide association of MASLD based on model 2. **B)** Comparison of the proteome-wide association between MASLD and BMI. T-statistics were obtained from logistic regression under model 2. Fitted linear regression model and Pearson’s correlation are shown. Axis labels show the number of cases and controls, or total samples used in the association analysis.

The 929 proteins associated with MASLD were further investigated for their co-regulation through network and enrichment analyses using the STRING database^14^. The proteins showed active interactions in the networks (Supplementary Figure 3) and were enriched in various biological pathways, diseases, and tissues, particular in immune system, lipoprotein, and visceral adipose and liver tissue-related pathways (Supplementary Table 7).

### Identification of proteins may be in the causal pathways of MASLD

For causal inference, we conducted MR analyses to distinguish proteins that cause MASLD from those that are affected by the disease or other biases. We used cis-pQTL data from GWAS of individual protein in UK Biobank for the MR analyses. We identified 824 out of the 929 MASLD-associated proteins having at least one cis-pQTL.

To increase the power of GWAS summary statistics for MASLD, we utilized multiple sources including three previous studies (GWAS_Ghodsian_,^16^ GWAS_Finngen_,^17^ and GWAS_MVP_^46^) and three in-house sources: GWAS_UKB_, GWAS_meta_, and GWAS_meta+MVP_ (Methods; Supplementary Table 8; Supplementary Figure 4). GWAS_meta_ included 19,477 MASLD cases and 886,736 controls, and GWAS_meta+MVP_ included 109,885 MASLD or proxy (unexplained chronic ALT elevation) cases and 1,014,923 controls, as the largest MASLD GWAS up to date. Our in-house GWAS identified five unique loci which were reported to be genome-wide significantly associated with MASLD for the first time (*p*<5×10^−8^; Table 2; Supplementary Figure 5). They included four loci based on GWAS_meta_ (*APP, CYP7A1/UBXN2B, RPL7P6/CAPZA3, and ZNF737*), and two loci based on GWAS_meta+MVP_ (*PEMT*, and *ZNF737*). Table 2 demonstrates the directional consistency of the associations across the various GWASs.

**Table 2.**
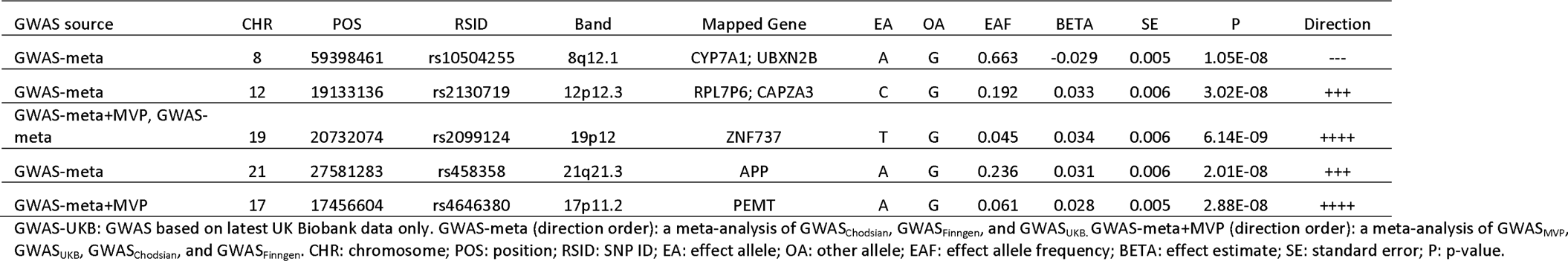
Genetic loci (top SNPs) firstly reported to be genome-wide significantly associated with MASLD based on in-house GWAS of MASLD.

Our MR analysis of the 824 plasma proteins and MASLD GWAS sources identified the genetic predisposition of nine proteins associated with MASLD, directionally consistent with the findings in the observational study. It included plasma APOE, CD33, GRHPR, HMOX2, IL1RN, KRT18, and SORT1 with a higher risk of MASLD and plasma NCAN and SCG3 with a lower risk of MASLD (*p*<1.2×10^−4^; Figure 3A, Supplementary Table 9). Among them, CD33, GRHPR, HMOX2, and SCG3 are newly identified proteins associated with MASLD or NAFLD. The association was robust using various MR methods, including simple mode, weighted mode, weighted median, MR Egger, and/or MR-PRESSO^25,26^ (Figure 3B). Further MR-Egger intercept test, MR-Egger regression and contamination mixture method^27^ also revealed that pleiotropy, and heterogeneity did not influence the associations between the nine significant proteins and MASLD (Supplementary Table 9). Colocalization analysis suggested that the genetic associations with plasma APOE, KRT18, and IL1RN were due to the same genetic variants associated with MASLD^30^ (posterior probability ≥ 0.7; Supplementary Figure 6). Comparing MR results in independent MASLD GWAS sources (GWAS_MVP_, GWAS_Finngen_, and GWAS_UKB_) revealed directionally consistent and significant associations with MASLD for APOE, IL1RN, NCAN, SORT1, and a novel target, SCG3 (*p*<0.05; Figure 3A). Further MR analyses using previous Somalogic proteome data in an independent population^20^ replicated the association of APOE, IL1RN, NCAN, and SCG3 with MASLD (*p*<8.3×10^−3^; Figure 3C; Supplementary Table 10).

**Figure 3.**
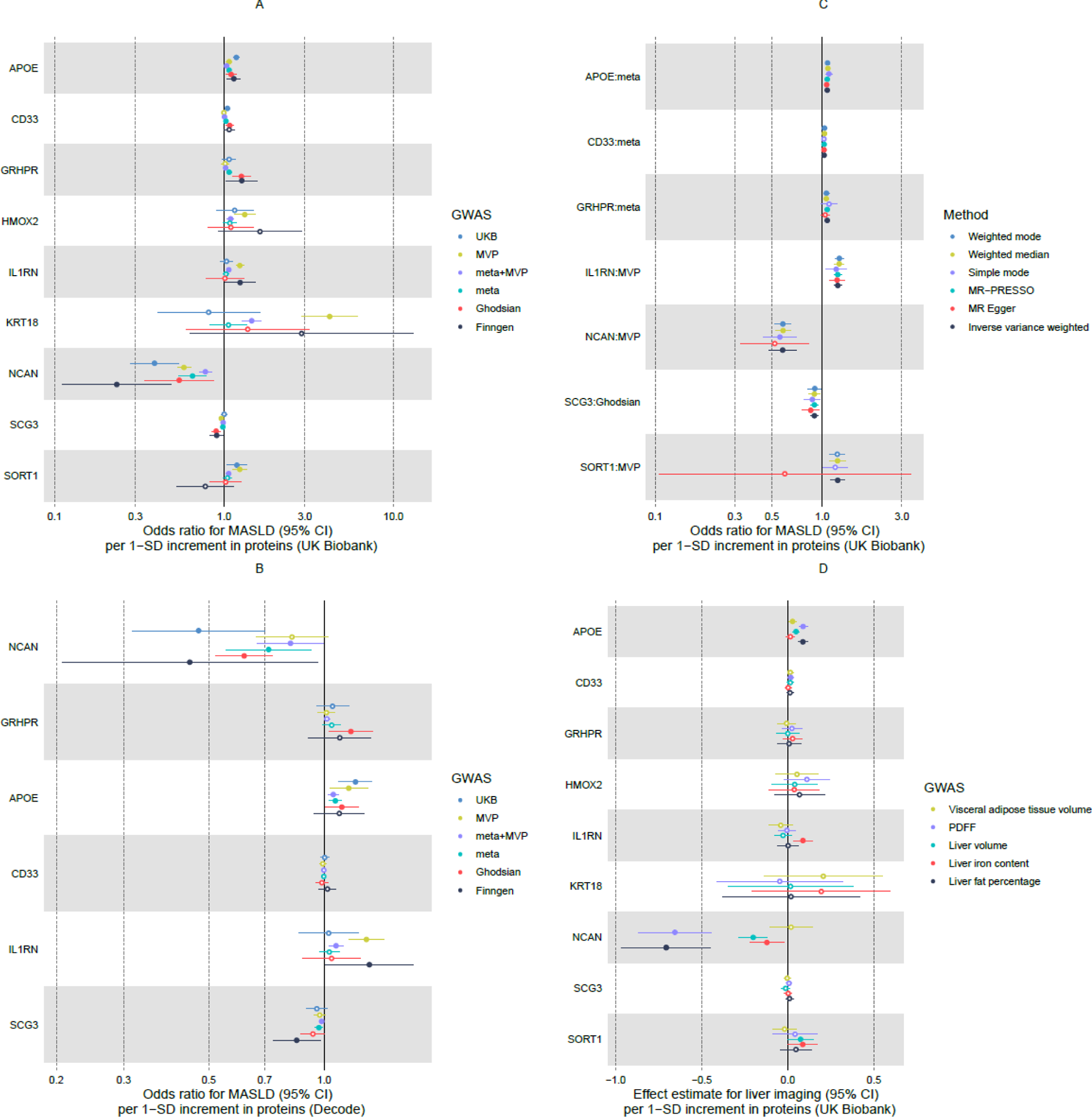
Association of proteins and MASLD by MR analysis. **A)** Association of proteins and MASLD by MR analysis through different MASLD GWAS sources. Proteins significantly associated with at least one MASLD GWAS source are shown (*p*<1.2×10^−4^). **B)** Association of proteins and MASLD by different MR methods, if applicable. The figure shows the MASLD GWAS source underlying the most significant result by inverse variance weighted MR. **C)** Association of proteins measured by Somalogic in decode and MASLD by MR analysis through different MASLD GWAS sources. Six proteins are available and shown. **D)** Association of proteins and liver imaging variables. Solid point indicates *p-value* less than 0.05. Hollow point indicates *p-value* not less than 0.05. Detailed data are presented in Supplementary Table 9, 10 and 13.

We hypothesized that proteins observationally associated with MASLD are more likely to be on the causal pathway of the disease if their encoding genes are also associated with MASLD. We found that 11 of the 929 MASLD-associated proteins with at least one MASLD-associated single-nucleotide polymorphism (SNP; *p*<5×10^−8^) in or near their encoding genes. In addition to the proteins also identified in the MR above (APOE, IL1RN, and NCAN) these included AHCY, ANPEP, APOC1, FCGR2B, KRT8, LPL, ORM1, and RBKS (Supplementary Table 11). Among them, AHCY, FCGR2B, ORM1, and RBKS were identified on their plasma protein levels with MASLD or NAFLD for the first time.

By combining observational and genetic analyses, we identified 17 unique proteins that may play a role in the causal pathway of MASLD, including eight novel MASLD-associated plasma proteins (AHCY, CD33, FCGR2B, GRHPR, HMOX2, ORM1, RBKS, and SCG3) and nine known MASLD-associated proteins (ANPEP, APOC1, APOE, IL1RN, KRT18, KRT8, LPL, NCAN, and SORT1).

### Integration of the 17 candidate causal MASLD proteins with multi-dimensional data

To further understand the causal pathways implicated by the 17 MASLD-associated proteins, we conducted further cross-omics annotation, which are summarized in Figure 1 and Table 3.

**Table 3.**
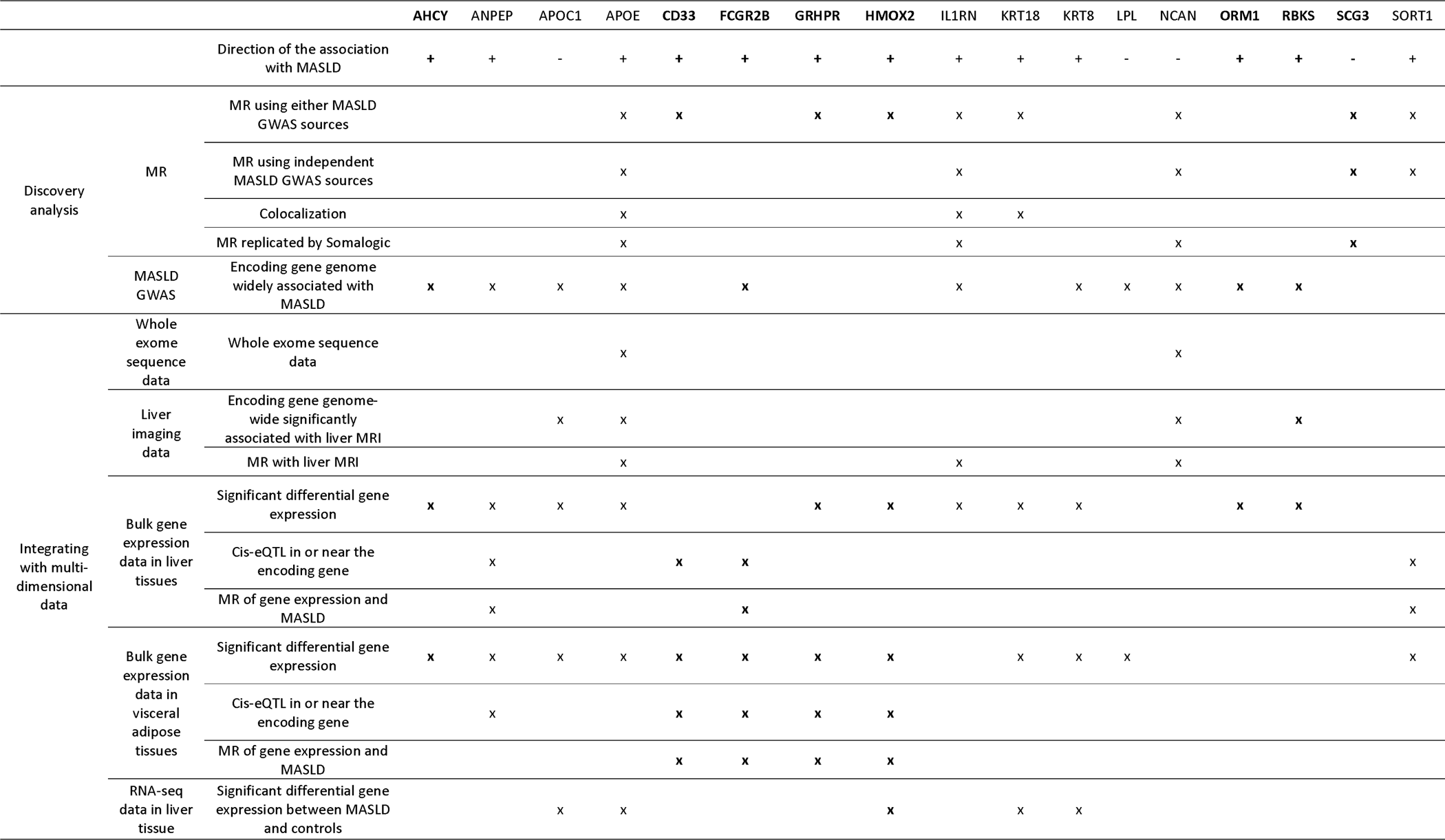
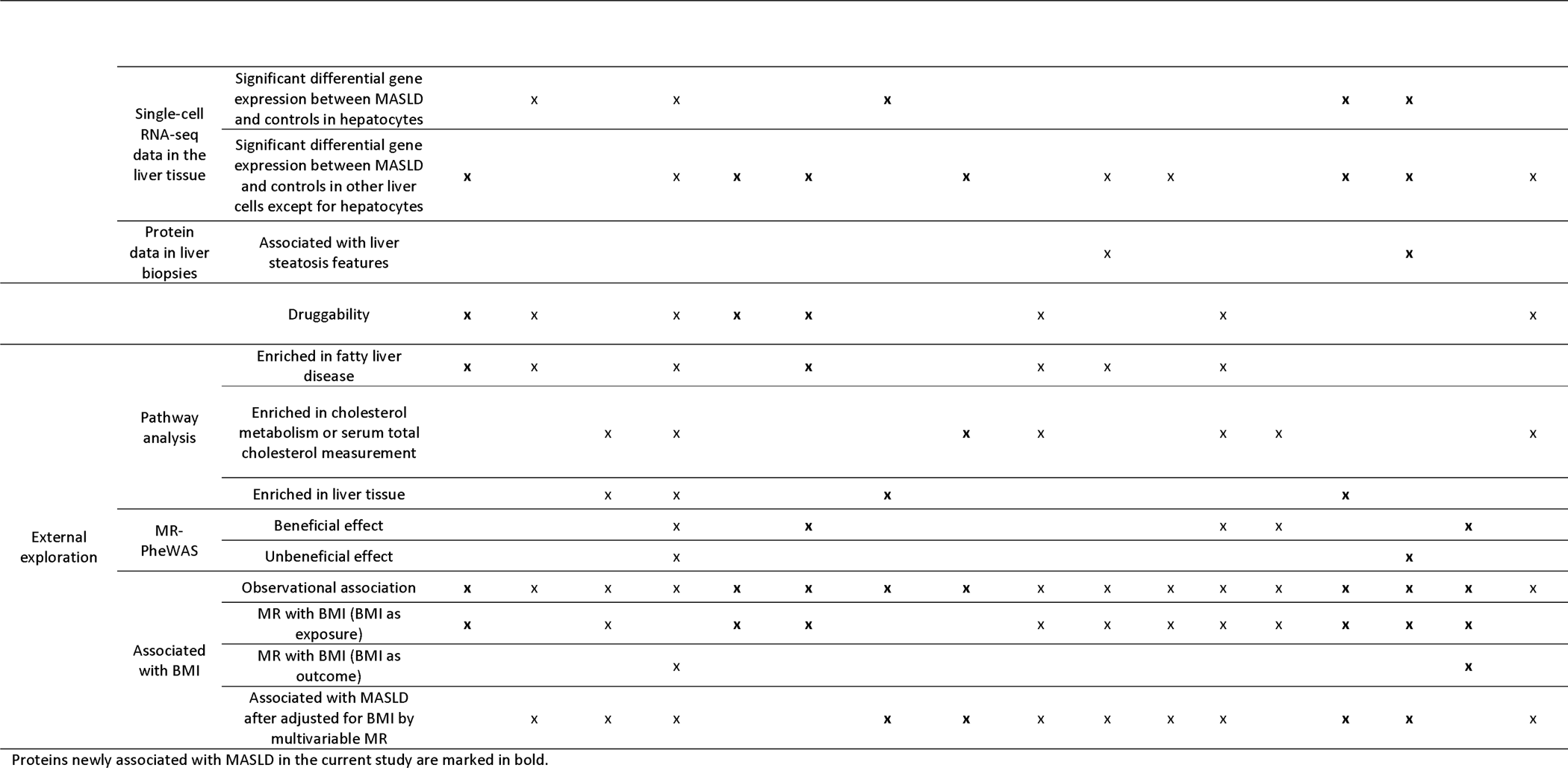
Summary of the cross-omics annotation of the 17 candidate causal MASLD proteins.

#### Integrating with exome sequencing data

We tested whether genetic variants in the exome sequencing data from UK Biobank (n∼470K, *p*<5×10^−8^; Methods) were associated with both MASLD and the candidate causal proteins in-cis and directionally consistent with the observational findings. We focused on variants with moderate to high impact on gene function^31^. We found that the missense variant at 19:19219115 (minor allele frequency, MAF = 0.075) was associated with plasma NCAN and MASLD, and the common missense variants at 19:44905910 (MAF = 0.36) and 19:44908684 (MAF=0.15) were associated with plasma APOE and MASLD (Supplementary Figure 7; Supplementary Table 12).

#### Integrating with liver imaging data

We then explored various evidence from liver imaging for the 17 candidate causal proteins. We studied MRI-PDFF, liver volume, liver iron content, liver fat percentage, and visceral adipose tissue volume, assessed by liver MRI in UK Biobank. We found that APOC1, APOE, NCAN, and RBKS have at least one SNP, in or near their encoding genes (+/−500kb), genome-wide significantly associated with PDFF and liver volume, and NCAN and RBKS have at least one SNP genome-wide significantly associated with liver fat percentage (*p*<5×10^−8^; Supplementary Figure 8). MR analyses showed that genetically determined plasma APOE and NCAN levels were associated with PDFF, liver fat percentage, and liver volume, whereas genetically determined IL1RN levels were associated with liver iron content (*p*<3.3×10^−3^, Figure 3D; Supplementary Table 13).

#### Integrating with gene expression data in the liver and visceral adipose tissue

We further investigated the 17 plasma proteins with gene expression data to understand whether these candidate causal proteins might originate from the liver. We found that the encoding genes of AHCY, ANPEP, APOC1, APOE, GRHPR, HMOX2, IL1RN, KRT18, KRT8, ORM1, and RBKS were highly expressed in liver, and the encoding genes of AHCY, ANPEP, APOC1, APOE, CD33, FCGR2B, GRHPR, HMOX2, KRT18, KRT8, LPL, SGSH, and SORT1 were highly expressed in visceral adipose tissue (Supplementary Figure 9A). The up-regulated differential expression of these encoding genes was enriched in liver and visceral adipose tissues than other tissues (p<0.05/54; Supplementary Figure 9B). We identified ANPEP, CD33, FCGR2B, and SORT1 having at least one cis-eQTL for liver tissue in or near their encoding genes, and ANPEP, CD33, FCGR2B, GRHPR, and HMOX2 having at least one cis-eQTL for visceral adipose tissue (*p*<5×10^−8^; Supplementary Table 14). MR analyses show that the genetically determined expression levels of *ANPEP, FCGR2B* and *SORT1* in liver, and the genetically determined expression levels of *CD33*, *FCGR2B*, *GRHPR,* and *HMOX2* in visceral adipose tissue associated with MASLD, directionally consistent with the association between plasma proteins and MASLD (Supplementary Table 15). Using bulk RNA-seq data of liver biopsies from 206 MASLD patients and 10 controls^34^, we found that expression levels of *APOC1*, *APOE*, *HMOX2*, *KRT18* and *KRT8* were significantly different in the liver tissues of MASLD patients and non-MASLD controls, and in the consistent direction as shown in our observational findings (*p*<3.3×10^−3^, Supplementary table 16).

#### Integrating with snRNA-seq and scRNA-seq data

A further exploration on liver tissue cell-type-based differential gene expression of the encoding genes of the 17 candidate causal MASLD proteins was performed, combining snRNA-seq and scRNA-seq data from human liver biopsies from six previous publications^36–42^ (Supplementary Figure 10). We found that *ANPEP, APOC1, APOE, GRHPR, ORM1*, and *RBKS* were differentially expressed in hepatocytes than other genes in hepatocytes. *ANPEP, APOE, GRHPR, ORM1,* and *RBKS* expression levels were significantly (*p*<1.0×10^−3^) up-regulated in non-alcoholic fatty liver (NAFL)/NASH patients^39^, directionally consistent with our observational study in plasma. *APOE, KRT8, KRT18, ORM1, RBKS*, and *SORT1* were differentially expressed in cholangiocytes and up-regulated significantly in MASLD patients^38^. *AHCY, CD33*, and *FCGR2B* were differentially expressed in myeloid cells, and *HMOX2* was differentially expressed in T-lymphocytes, and up-regulated significantly in MASLD patients^38^.

#### Integrating with protein data in liver biopsy

We next associated the proteins with histological hepatic steatosis features using human liver biopsies from 36 women undergoing elective surgery (Methods). Eight candidate causal proteins (AHCY, ANPEP, APOC1, APOE, GRHPR, KRT8, KRT18, and RBKS) were available amongst the proteins measured in the liver biopsies using mass spectrometry. We identified KRT18 and RBKS levels in these liver biopsies were positively significantly associated with percentage of steatosis (*p*<8.3×10^−3^; Supplementary Table 17).

### Pathway enrichment analysis

We performed enrichment analyses of the 17 candidate causal proteins to understand biological mechanisms of MASLD (Supplementary Table 18)^43,47^. The top pathway enriched was cholesterol metabolism (APOC1, APOE, LPL, SORT1 and HMOX2), followed by extrinsic apoptotic signalling pathway (KRT8, KRT18, and SORT1) and regulation of interleukin-1 beta production (ANPEP, CD33, FCGR2B, HMOX2, LPL, and ORM1). The top three diseases enriched were alcoholic liver diseases (IL1RN, KRT8, KRT18, LPL, and NCAN), fatty liver disease (AHCY, ANPEP, APOE, FCGR2B, IL1RN, KRT18, and LPL), and serum total cholesterol measurement (APOC1, APOE, IL1RN, LPL, NCAN, and SORT1). APOC1, APOE, GRHPR and ORM1 were enriched in liver tissue and APOC1, IL1RN, and ORM1 were enriched in human liver cancer cell line (Hep-G2).

### PheMR

We performed a PheMR to explore potential effects of the 17 candidate causal proteins on other diseases beyond MASLD (*p*<1.0×10^−5^, Supplementary Figure 11, Supplementary Table 19). The PheMR results show that for five proteins in the causal pathway, modifying their plasma levels for lowering MASLD risks also reduces the risk of other diseases, including obesity and arthritis with APOE, depressive episode and ulcerative colitis with FCGR2B, acute myocardial infarction with LPL, type 2 diabetes with NCAN, and tympanic membrane disorders with SCG3. However, modifying the plasma APOE levels for MASLD may increase the risk of neurological degenerative disorders, such as Alzheimer’s disease, vascular dementia, cognitive disorders, and delirium, whereas modifying plasma RBKS levels may increase the risk of iron deficiency anaemia.

### Druggable analysis

To leverage the 17 potential causal proteins to therapeutic avenues for MASLD, we investigated their potential druggability through OpenTargets^48^ and Therapeutic Target Database^49^. APOC1, APOE, IL1RN, LPL, NCAN, ORM1, and SCG3 are secreted proteins which are easier accessible sources of therapeutics. We also identified nine unique drugs targeting five proteins, including ANPEP (Tosedostat/IP10C8), APOE (AEM-28), CD33 (Gemtuzumab ozogamicin/Lintuzumab/M195/AVE-9633), FCGR2B (Obexelimab), and LPL (Clofibrate).

### BMI in the causal association of proteins and MASLD

We further explored the role of BMI, the most important risk factor of MASLD, in the association between the candidate causal proteins and MASLD. We found that genetically determined BMI was associated with AHCY, APOC1, CD33, FCGR2B, IL1RN, KRT18, KRT8, LPL, NCAN, ORM1, and SCG3 (Figure 4A); while genetically determined APOE and SCG3 were associated with BMI (p<3.3×10^−3^; Supplementary Table 20; Figure 4B). Multivariable MR analysis showed that after adjusting for the causal effect of BMI on MASLD, genetically determined plasma ANPEP, APOC1, APOE, GRHPR, HMOX2, IL1RN, KRT18, KRT8, LPL, ORM1, RBKS, and SORT1 were still significantly associated with MASLD (p<3.3×10^−3^; Supplementary Table 21; Figure 4C). We did not detect significance between genetic determined plasma AHCY, CD33, FCGR2B, NCAN, and SCG3 with MASLD after adjusting for the causal effect of BMI on MASLD (Figure 4D).

**Figure 4.**
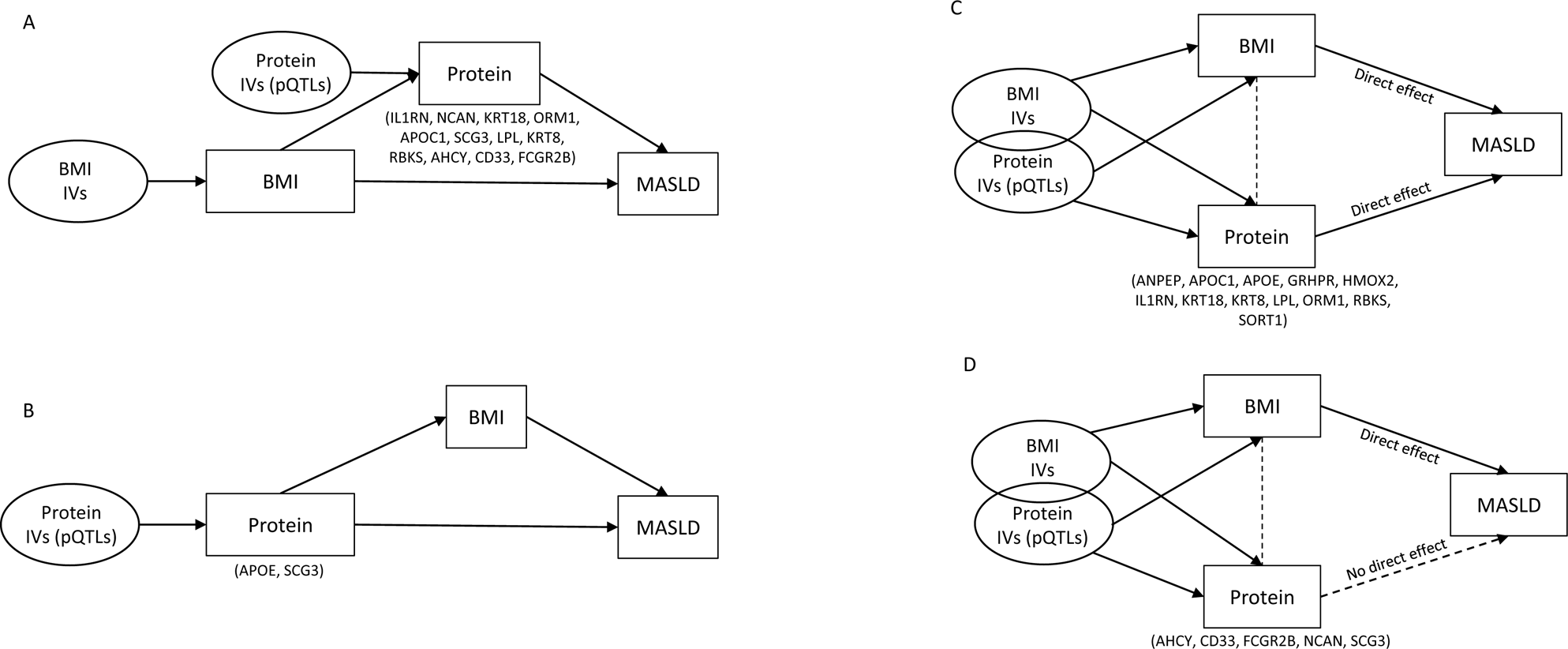
Role of BMI in the association of candidate causal proteins and MASLD. **A)** Proteins as mediators in the causal association of BMI to MASLD. **B)** BMI as a mediator in the causal association of proteins to MASLD. **C)** Proteins directly associated with MASLD after adjusted for the causal effect of BMI by multivariable MR. **D)** Proteins not directly associated with MASLD after adjusted for the causal effect of BMI by multivariable MR. IVs: instrumental variables. Arrows indicate direction from MR analysis, or knowledge (i.e., IVs causes exposures, and BMI causes MASLD).

## Discussion

Using large-scale genomic and proteomic data, we identified 929 (31.6%) plasma proteins associated with MASLD. Of these proteins, 17 were potentially involved in the causal pathway of MASLD (Table 3). By adopting a plasma proteome-first approach and integrating with human genetics, transcriptomics, liver imaging, and biopsy proteomics, we found four novel protein/genetic targets of MASLD, namely CD33, GRHPR, HMOX2, and SCG3. Additionally, our study provided evidence supporting AHCY, FCGR2B, ORM1, and RBKS on MASLD from proteins, and validated previously known MASLD targets by MR, including ANPEP, APOC1, APOE, IL1RN, KRT18, KRT8, LPL, NCAN, and SORT1.

We used the largest proteomics dataset currently available to discover proteins associated with MASLD. Comparing to a recent study by Sveinbjornsson et al.^50^, which found one protein associated with NAFLD by MR, we identified nine proteins potentially on the causal pathway to MASLD using MR. Our GWAS helped to identify five novel MASLD loci (*APP, CYP7A1, PEMT, RPL7P6/CAPZA3, and ZNF737*), of which the expression of *CYP7A1* and *PEMT* were reported previously with NAFLD^51,52^. These novel loci were associated (*p*<5×10^−8^) with other traits such as Alzheimer’s disease (*APP*), height (*APP*, *PEMT*, and *RPL7P6/CAPZA3*), BMI and lung function (*RPL7P6/CAPZA3*), waist-hip index and coronary artery disease (*PEMT*), lipid levels (*APP*, *CYP7A1*, *PEMT*, and *ZNF737*) and gallstones, cholelithiasis, liver enzymes, Insulin-like growth factor 1 levels and bilirubin (*CYP7A1*) ^53^.

We have identified CD33, GRHPR, HMOX2, and SCG3 as novel targets of MASLD. Among them, GRHPR and HMOX2 had independent effects on MASLD beyond obesity/BMI. Glyoxylate and hydroxypyruvate reductase (GRHPR) reduces glyoxylate into less reactive glycolate and is mainly present in the liver. Previous studies have reported impaired glyoxylate detoxification in MASLD^54^, which may explain the causal association of plasma GRHPR and MASLD beyond obesity. Heme oxygenase 2 (HMOX2) is a genetically distinct isozymes of heme oxygenase 1 (HMOX1), and previous studies have reported that increased expression of *HMOX1* in human liver biopsies reflects the severity of MASLD^55^. The colocalization of *HMOX2* expression in tibial artery and muscle with blood insulin-like growth factor (IGF) 1 levels suggests potential pathways through artery and/or muscle to MASLD. Secretogranin III (SCG3) is a secreted protein which has been reported previously to regulate IGF transport and insulin secretion^56^ and is associated with BMI^53,57^. Our study provides robust causal evidence of the effect of plasma SCG3 on MASLD, which was disappeared when adjusted for the causal effect of BMI on MASLD, suggesting the importance of obesity in the association of SCG3 and MASLD. Few previous studies focused on the association of CD33 and MASLD, though approved drugs of CD33 are available.

Moreover, we deduced the causal association of plasma AHCY, FCGR2B, ORM1, and RBKS with MASLD involving MASLD GWAS studies. These targets were not previously highlighted in the GWAS, potentially because GWAS tend to map target genes based on the top genetic loci/SNP, ignoring secondary associations with neighbouring genes. Our study’s pipeline caught this missing information by looking up the full set of genes mapped in any genome-wide significant SNPs of MASLD, validated by their encoding proteins associated with MASLD. Ribokinase (RBKS) is one of the genes which was in a known MASLD region but over-shadowed by the primary association with *GCKR*. Our study found a consistent association of liver RBKS levels with steatosis features in liver biopsies, along with significant differential gene expression in liver tissue, mainly hepatocytes and cholangiocytes. This finding suggests that carbohydrate metabolism, the main pathway of RBKS, in the liver may alter the incidence of MASLD. Orosomucoid 1 (ORM1) is another target of MASLD, which was masked by *AKNA*. It is a secreted protein and transports proteins in the blood stream but is also highly expressed in human hepatocytes and cholangiocytes^38,39^. Its isoform ORM2 regulates de novo lipogenesis, one of the most important mechanisms of MASLD, in the mouse liver, which may indicate the potential causal association of ORM1 and MASLD^58,59^. A few studies have reported the potential association of Adenosylhomocysteinase (AHCY)^60^ and Fc gamma receptor IIb (FCGR2B) with MASLD^61,62^. Our study found that their causal association with MASLD disappeared after adjusting for the causal effect of BMI.

Our study provides causal evidence for previous MASLD targets, including ANPEP, APOC1, APOE, IL1RN, KRT18, KRT8, LPL, NCAN, and SORT1^63–69^. Notably, there have been approved drugs targeting ANPEP, APOE, LPL and SORT1^48,49^, though their effect on liver diseases is still not clear. We also provided various evidence for these targets, including gene expression in liver or single cells, liver imaging, liver biopsies, and potential beneficial or unbeneficial effects to other diseases. Our study examined the association of these targets with BMI, showing that BMI has causal mediating effect on the association of APOE to MASLD, and APOC1, IL1RN, KRT18, KRT8, and LPL have causal mediating effects on the association of BMI to MASLD. Additionally, we found that the causal association between NCAN and MASLD may be fully explained by the causal effect of BMI; while ANPEP, APOC1, IL1RN, KRT18, KRT8, LPL, and SORT1 have causal effects on MASLD beyond obesity. This suggests that, besides obesity, these proteins may be involved in other pathways relevant for MASLD that requires further exploration.

Despite being a comprehensive study identified many innovative findings of the targets for MASLD, this study has some limitations. Firstly, liver imaging was not available for all participants, which may have led to an underestimation of MASLD cases and increased the rate of false-negative results in MASLD associations. Secondly, the study relied on plasma proteins to identify potential therapeutic targets for MASLD, which may not necessarily reflect protein levels in liver tissue. However, this approach is informative, and partially validated by tissue data. Thirdly, the MASLD GWAS by Vujkovic M, et al used chronic ALT elevation as a proxy for MASLD, but their conclusion was well validated^70^ and our findings were generally consistent across cohorts (Figure 3). Fourthly, the shared genetic variation between MASLD and proteins was limited, which may be due to violations of the single causal variant assumption or the known high false-negative rate for colocalization^71^. Fifthly, when comparing snRNA-seq and scRNA-seq data between MASLD/NASH and non-MASLD groups, selection bias from limited participants may drive the conclusions.

Overall, our study underscores the immense potential of large-scale multi-omics in enhancing our understanding of complex diseases, such as MASLD in our study, and identifying potential targets for translational research. By leveraging these innovative technologies, we were able to shed new light on the pathophysiology of MASLD and uncover promising new avenues for therapeutic intervention.

## Author contributions

J.L., J.M.M.H. and C.M.v.D. conceived and designed the current study. J.L., T.G.R., L.C., S.H., and M.T., performed data analysis of UK Biobank. C.D. and R.M. performed analysis of proteins in the liver biopsy study. M.V., P.S.T., J.A.L., B.F.V, and K-M.C. contributed to the generation of MASLD data in MVP. C.A.P.M. and T.M. performed analysis of snRNA-seq and scRNA-seq data. J.F.C. contributes to clinician’s perspective. N.D., and J.W.T. contributed to the study design and participant recruitment for the liver biopsy study. J.L., S.H., T.G.R., C.D., C.A.P.M., C.M.v.D., and J.M.M.H. prepared the manuscript. All authors read, revised, and approved the manuscript.

## Data availability

UK Biobank data are publicly available to bona fide researchers upon application at http://www.ukbiobank.ac.uk/using-the-resource/. Publicly available summary statistics are obtained from https://gwas.mrcieu.ac.uk/, https://www.ebi.ac.uk/gwas/, https://www.gtexportal.org/home/datasets, and https://pan.ukbb.broadinstitute.org. Bulk liver RNA-seq data are available in Gene Expression Omnibus (GEO) under accession GSE135251. SnRNA-seq and scRNA-seq data are available in GEO under accession GSE189175, GSE185477, GSE136103, GSE212837, GSE189600, and GSE192740. Other source of data or web source were clarified in the methods. Relevant detailed results generated in this study were presented in supplementary tables. Clinical data with respect to the liver biopsy samples were assessable through contacting J.W.T.

## Acknowledgements

This research was conducted using data from UK Biobank, a major biomedical database (https://www.ukbiobank.ac.uk/) via application no. 53639 and 65851. We thank the participants, contributors, clinicians, and researchers for making data available for this study. We are grateful to the research & development leadership teams at the thirteen participating UKB-PPP member companies (Alnylam Pharmaceuticals, Amgen, AstraZeneca, Biogen, Bristol-Myers Squibb, Calico, Genentech, Glaxo Smith Klein, Janssen Pharmaceuticals, Novo Nordisk, Pfizer, Regeneron, and Takeda) for jointly funding the proteomics study in UK Biobank. We thank the team at Olink Proteomics for their consistent logistic support throughout the project. We thank Dipender PS Gill for his help in facilitating MVP collaboration. This research is based on data from the MVP, Office of Research and Development, Veterans Health Administration and was supported by award MVP000 (mvp003/028b). MVP MASLD data was generated with Department of Veterans Affairs (VA) funding support (I01-BX003362: K.M.C., P.S.T., and M.V.) with additional support from resources and facilities of the VA Informatics and Computing Infrastructure (VINCI), VA HSR RES 13-457. This publication does not represent the views of the Department of Veterans Affairs or the United States Government. M.V. acknowledges support for this work from the National Institutes of Health (NIH)/National Institute of Diabetes and Digestive and Kidney Diseases, grant R01 DK134575. J.L. is supported by a Novo Nordisk Postdoctoral Fellowship and N.D. through a Novo Nordisk clinical research training fellowship run in partnership with the University of Oxford. The work has been supported by the Medical Research Council (JWT) and by the Oxford NIHR Biomedical Research Centre (JWT).

## ABBREVIATIONS

MASLD: metabolic dysfunction-associated steatotic liver disease
NAFLD: non-alcoholic fatty liver disease
MASH: metabolic dysfunction-associated steatohepatitis
NASH: non-alcoholic steatohepatitis
MR: Mendelian randomization
MRI-PDFF: magnetic resonance imaging - estimated proton density fat fraction
ICD: International Classification of Diseases
LC-MS/MS: liquid chromatography-tandem mass spectrometry
MSD: Matrix Spectral Decomposition
GWAS: genome-wide association study
ALT: alanine aminotransferase
PheMR: phenome-wide Mendelian randomization study
pQTL: protein quantitative trait locus
eQTL: expression quantitative trait locus
BMI: body mass index
VEP: variant effect predictor
LOFTEE: loss-of-function transcript effect estimator
DEG: differentially expressed genes
snRNA-seq: single nucleus RNA sequence
scRNA-seq: single cell RNA sequence
age^2^: age squared
SNP: single-nucleotide polymorphism
NAFL: non-alcoholic fatty liver
GRHPR: Glyoxylate and hydroxypyruvate reductase
HMOX2: heme oxygenase 2
HMOX1: heme oxygenase 1
IGF: insulin-like growth factor
SCG3: secretogranin III
RBKS: ribokinase
ORM1: Orosomucoid 1
AHCY: adenosylhomocysteinase
FCGR2B: Fc gamma receptor IIb.

