## Supplementary Figure for "Profiling the genome and proteome of metabolic dysfunction-associated steatotic liver disease identifies potential therapeutic targets"

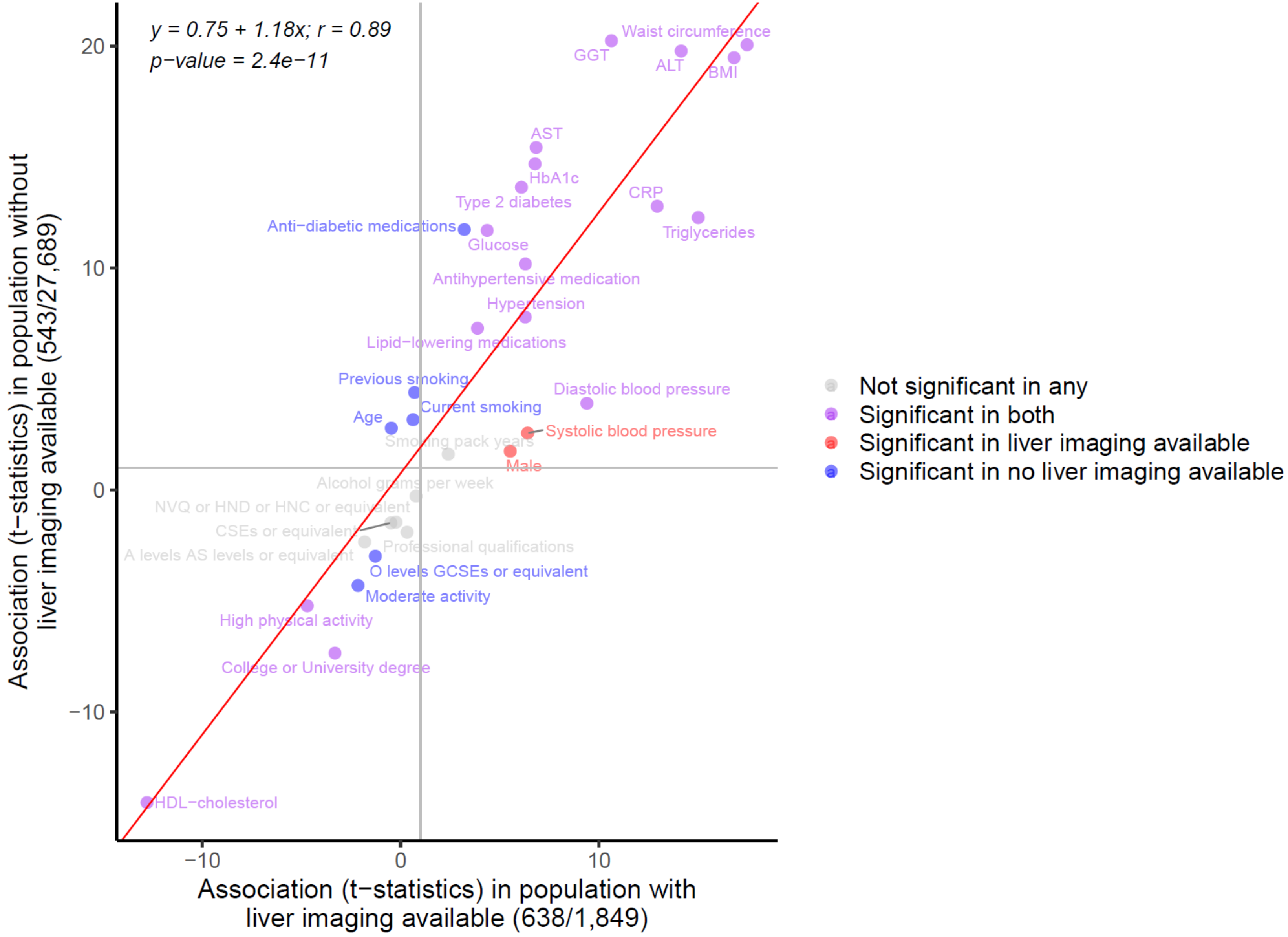


**Supplementary Figure 1 Comparison of the association patterns of MASLD and its common risk factors in the population with and without liver imaging available.**

T-statistics were obtained from logistic regression with age and sex adjusted. Fitted linear regression model and Pearson's correlation are shown. Axis labels show the number of MASLD cases and controls used in the association analysis.

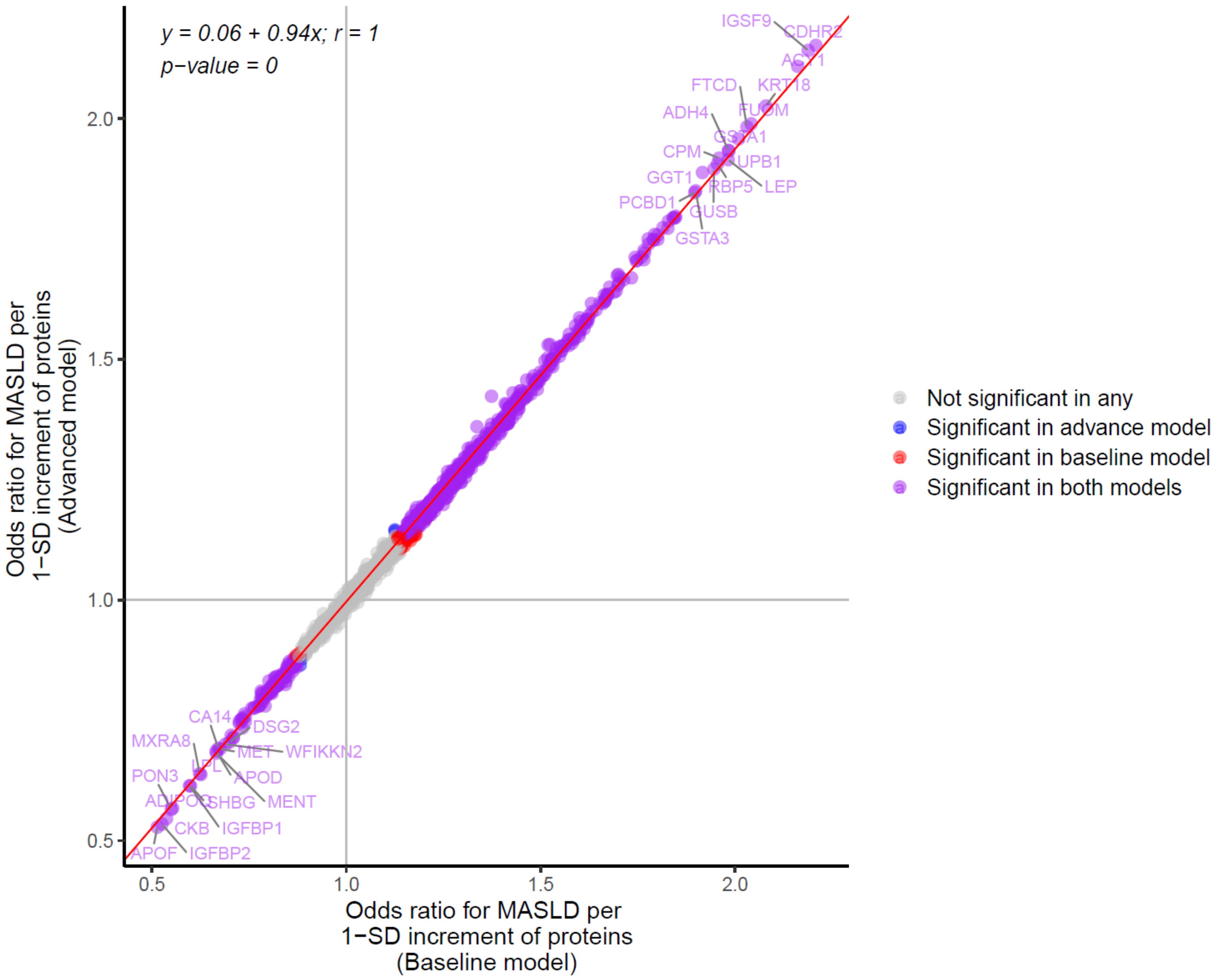


**Supplementary Figure 2 Comparison of the proteome-wide association of MASLD between the baseline model and advanced model.** Baseline model (model 1) adjusted for age, age squared (age2), sex, fasting time, and batch of proteomic measurement. Advanced model (model 2) additionally adjusted for common lifestyle factors, including smoking status, number of pack-years of smoking, alcohol consumption, education, and physical activity on top of model 1. Fitted linear regression model and Pearson's correlation are shown.

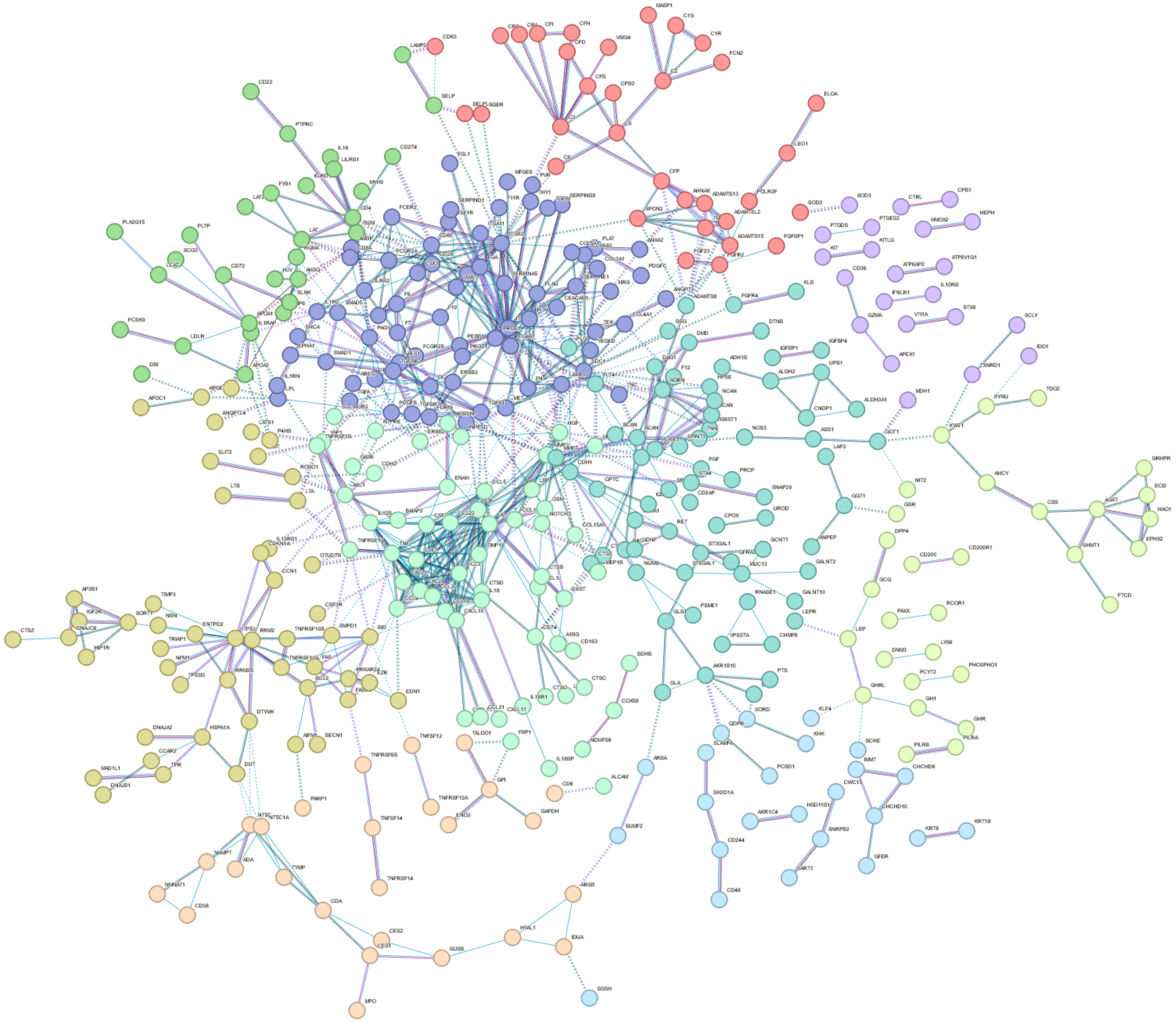

**Supplementary Figure 3 Protein-protein association network of MASLD-related proteins through STRING database.** 905 of the 929 MASLD-related proteins were identified in STRING database. Ten protein clusters are shown in different colours. Disconnected nodes in the network are hided. Edge colours: Blue: Association in Curated Databases; Rose: Experimental/Biochemical Data; Black: Co-Expression; Grey: the two proteins may have some sequence similarity; Dotted line: between clusters; solid line: between proteins within the same cluster.

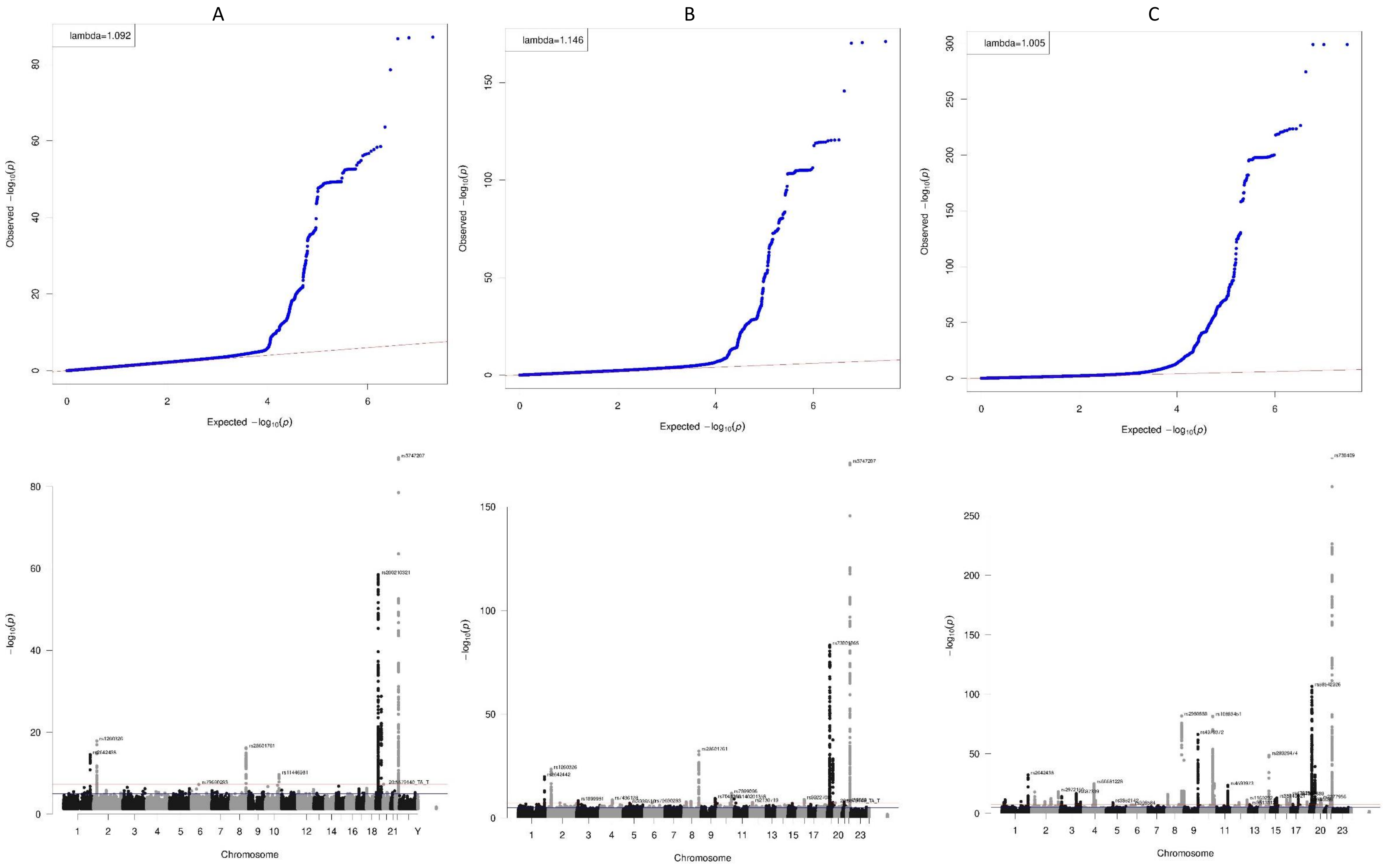


**Supplementary Figure 4 QQ plots and Manhattan plots of the three in-house GWAS of MASLD: $GWAS_{UKB}$ (A); $GWAS_{meta}$ (B); $GWAS_{meta+MVP}$ (C).**

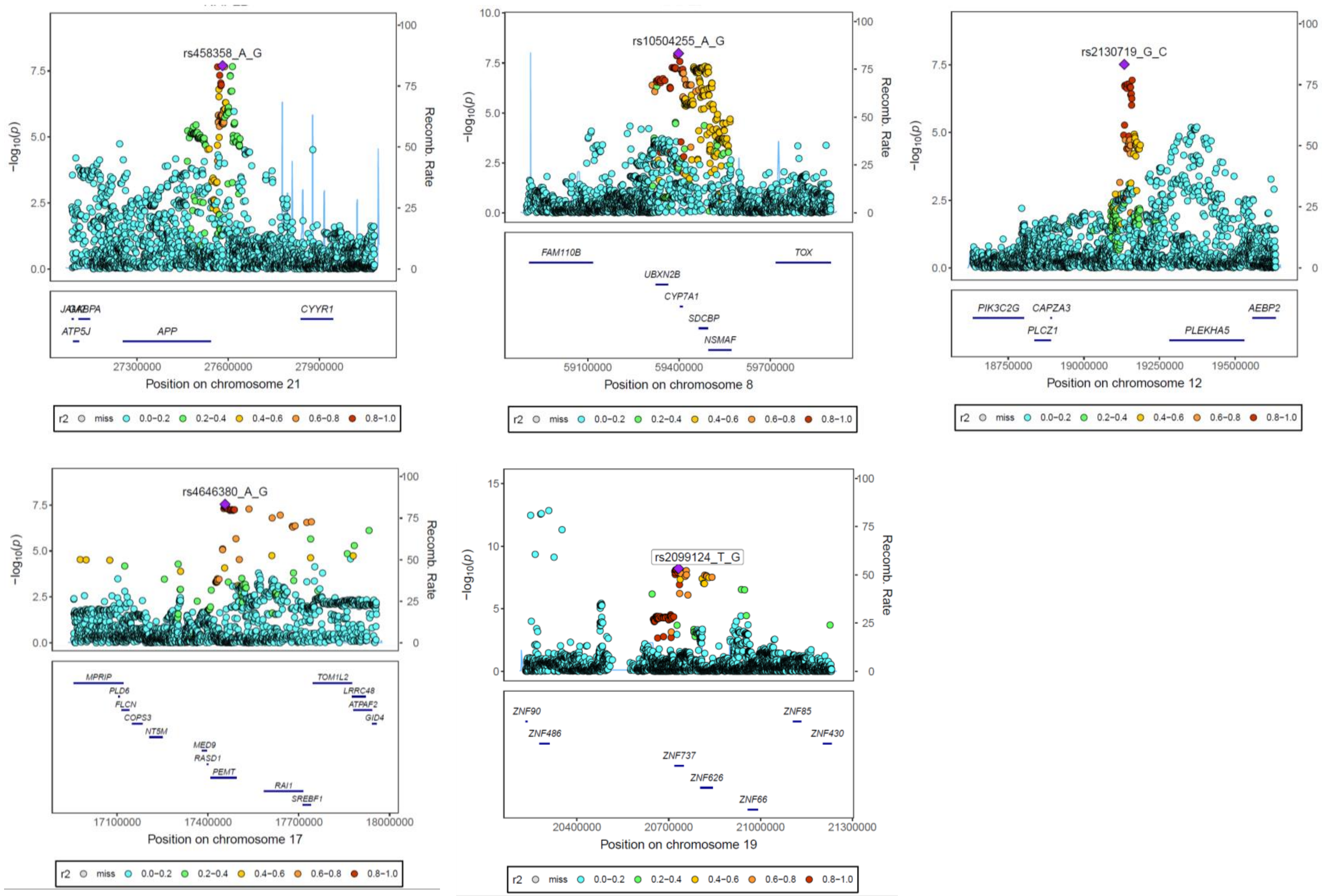


**Supplementary Figure 5 Locuszoom plots of the loci genome-widely associated with MASLD which were not reported previously**

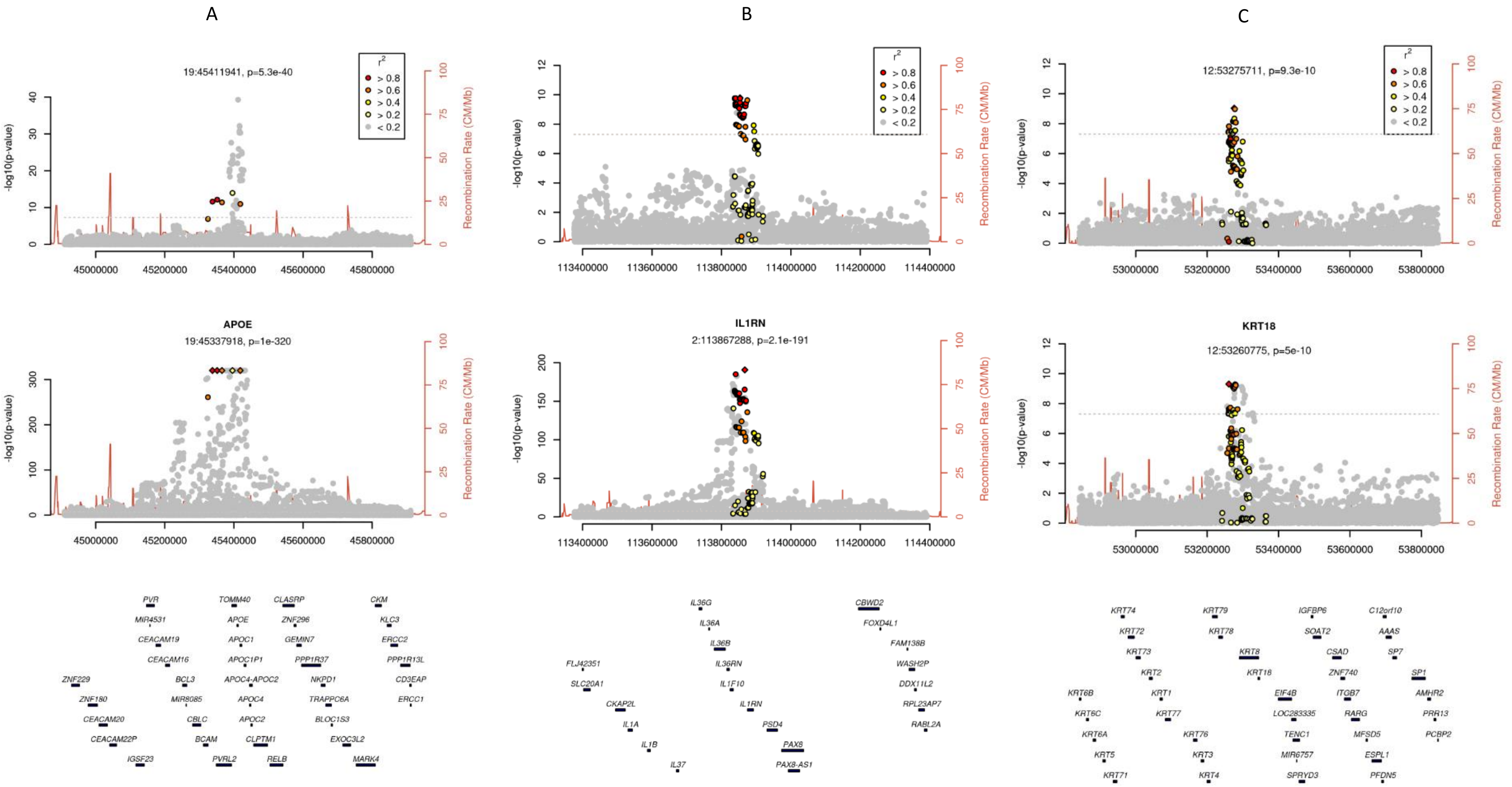


**Supplementary Figure 6 Locuszoom plots to illustrate evidence of genetic colocalization between proteins and MASLD.** The proteins with posterior probability ≥0.7 are shown. MASLD GWAS is based on $GWAS_{meta+MVP}$.

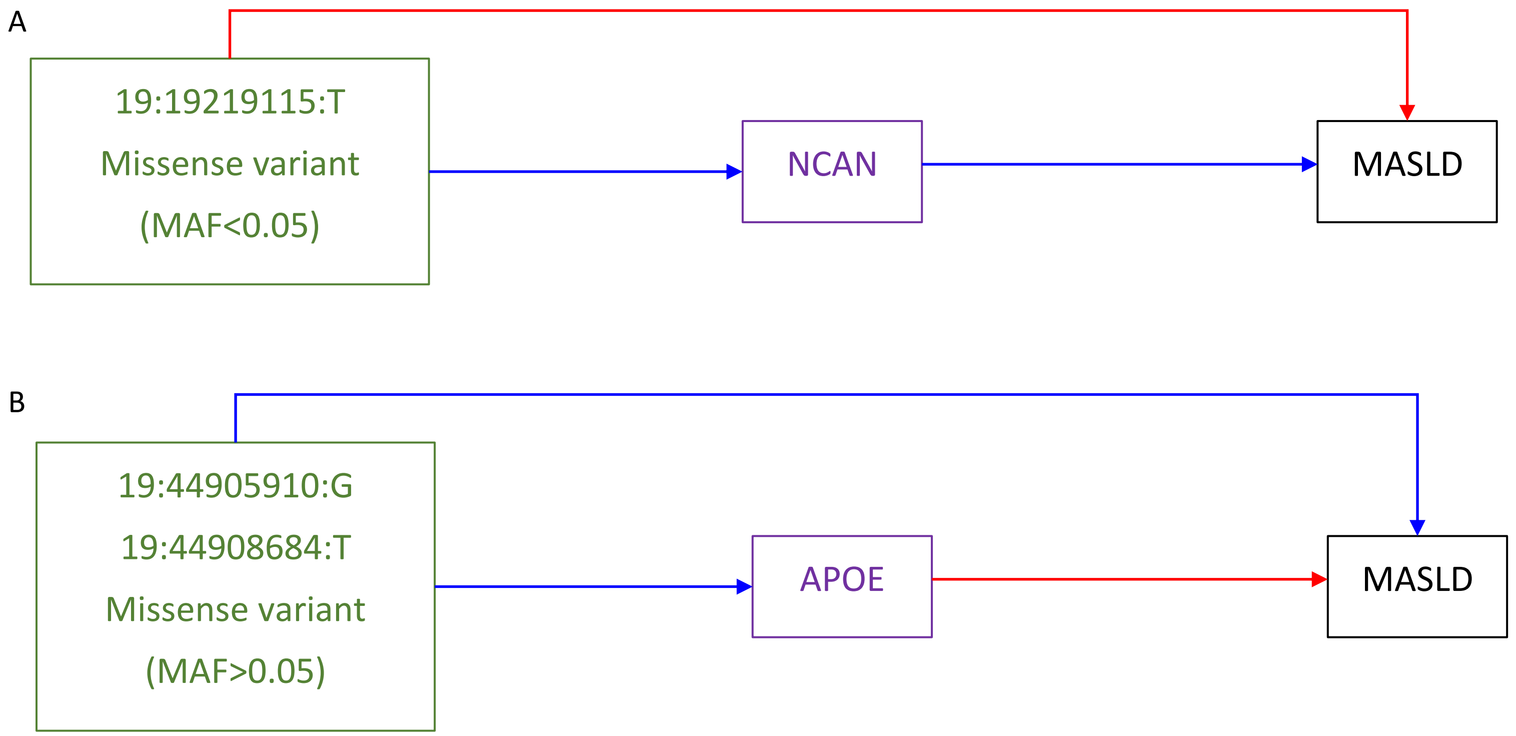


**Supplementary Figure 7 Significant pathways supported by exome sequencing data.** Green boxes: genetic variants shown as chromosome: position: effect allele. Purple boxes: plasma proteins. Red lines: positive association. Blue lines: negative association. Arrow: potential causal directions.

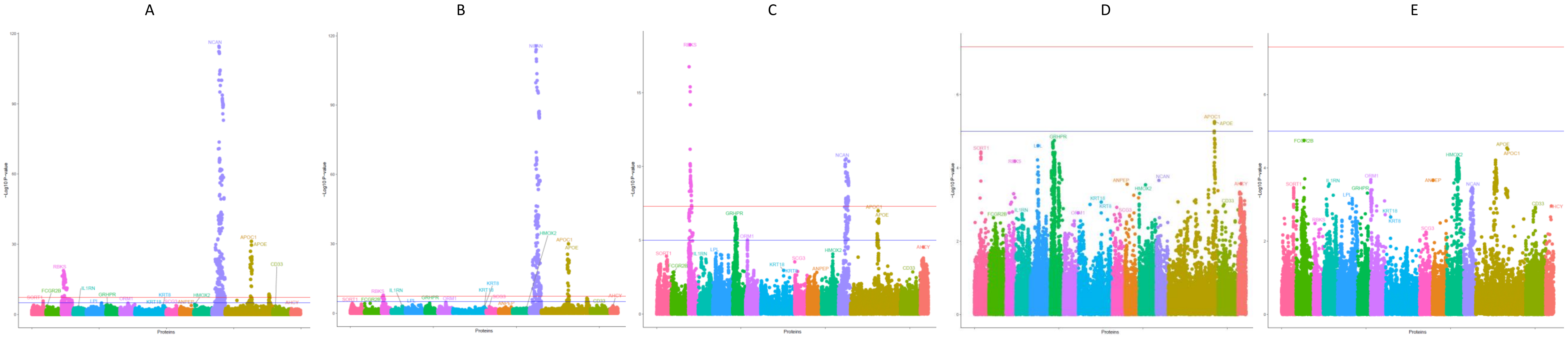


**Supplementary Figure 8 Associations of SNPs in/near the encoding genes of the 17 candidate causal proteins of MASLD and liver imaging variables: PDFF (A), liver fat percentage (B), liver volume (C), visceral adipose tissue volume (D), and liver iron content (E).** Red line: $p<5\times10^{-8}$; blue line: $p<1\times10^{-5}$.

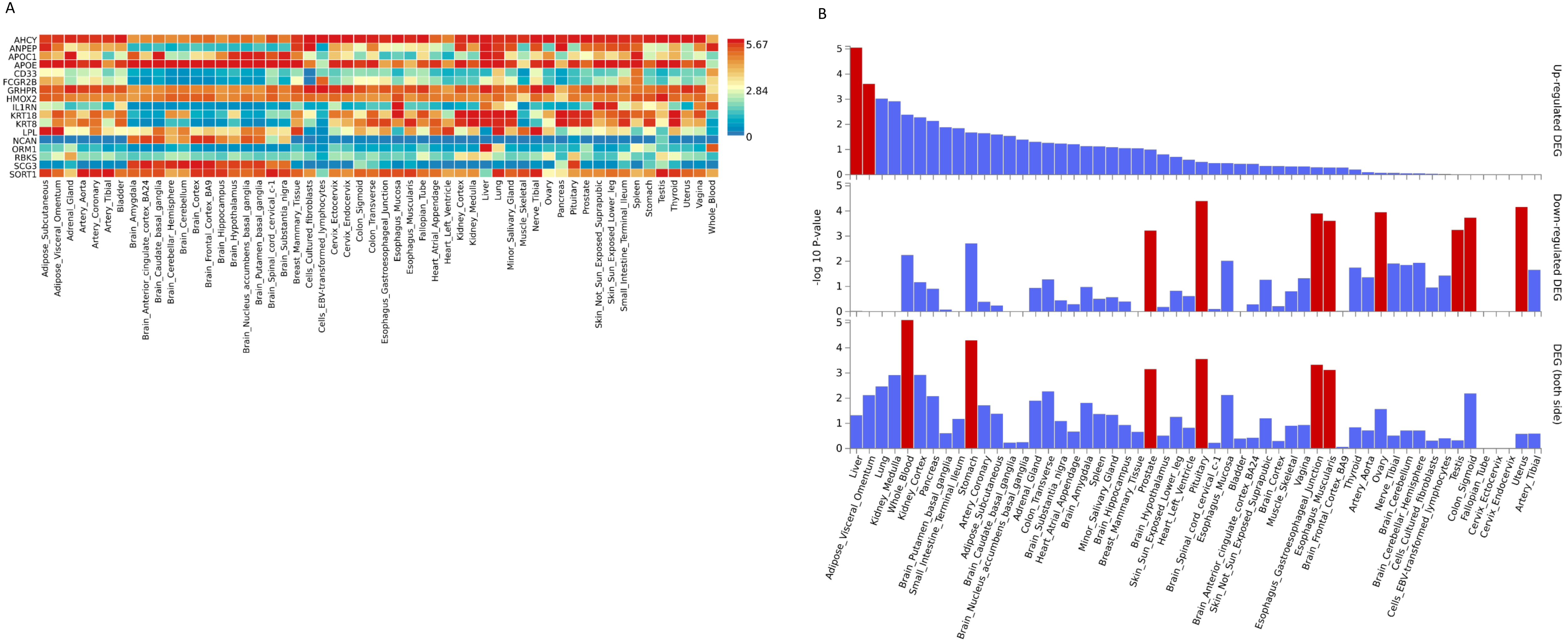


**Supplementary Figure 9 Gene expression levels (A) and tissue specificity (B) of the encoding genes of the 17 candidate MASLD proteins in different tissues.** The Figure was generated based on the default pipeline using FUMA with GTEx (v8) dataset in 54 tissue types (K Watanabe, et al. Nat Commun, 2019). The colors in figure A show the average expression per label (log2 transformed). Red color in figure B indicates significance.

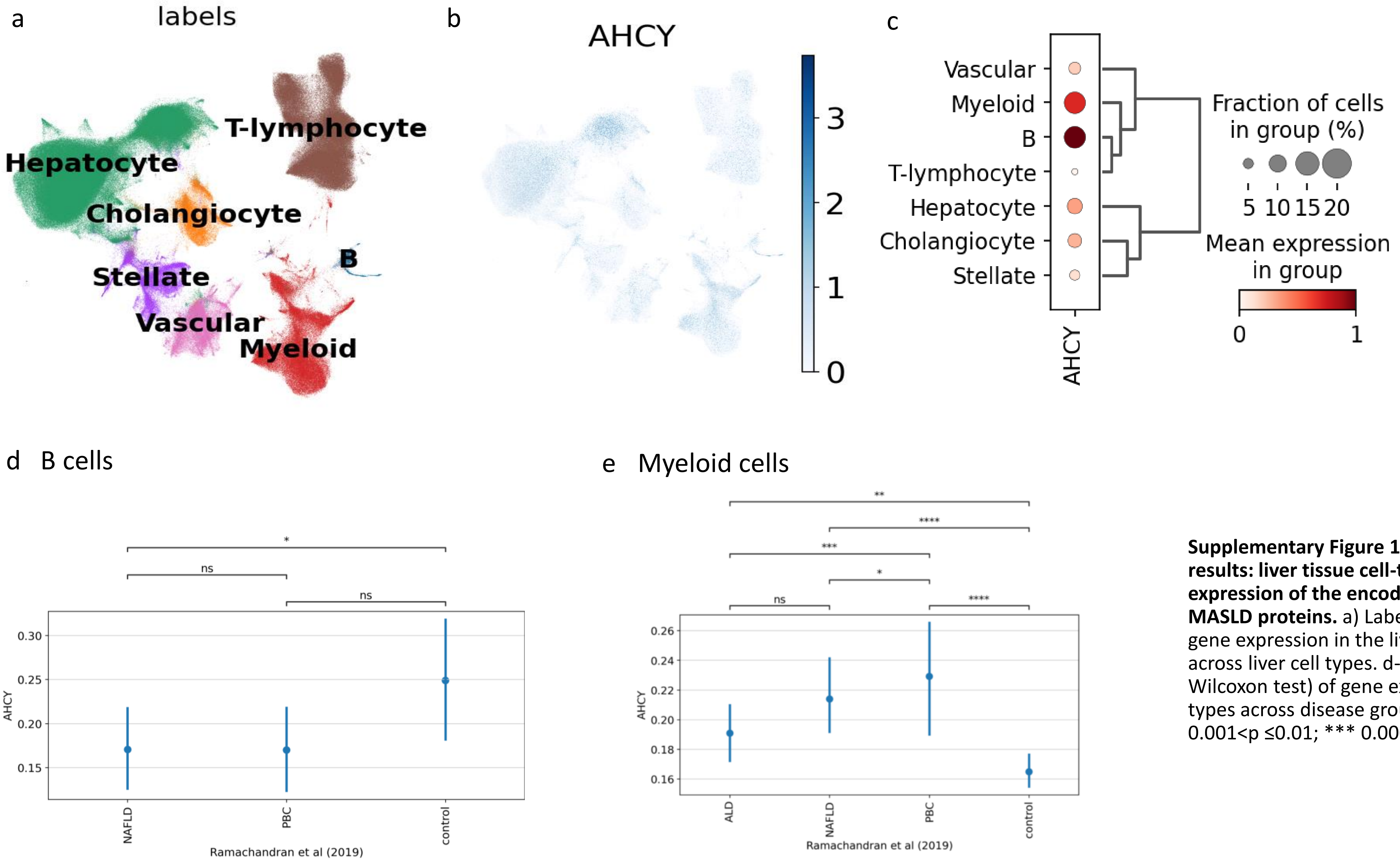


**Supplementary Figure 10 ScRNA-seq and snRNA-seq results: liver tissue cell-type-based differential gene expression of the encoding genes of the 17 candidate MASLD proteins.** a) Labels of UMAP. b) UMAP of the gene expression in the liver. c) Gene expression levels across liver cell types. d-f) Comparison (Mann-Whitney-Wilcoxon test) of gene expression levels in single cell types across disease groups. Ns, p≤1; * 0.01<p ≤0.05; ** 0.001<p ≤0.01; *** 0.0001<p ≤0.001; **** p ≤0.0001.

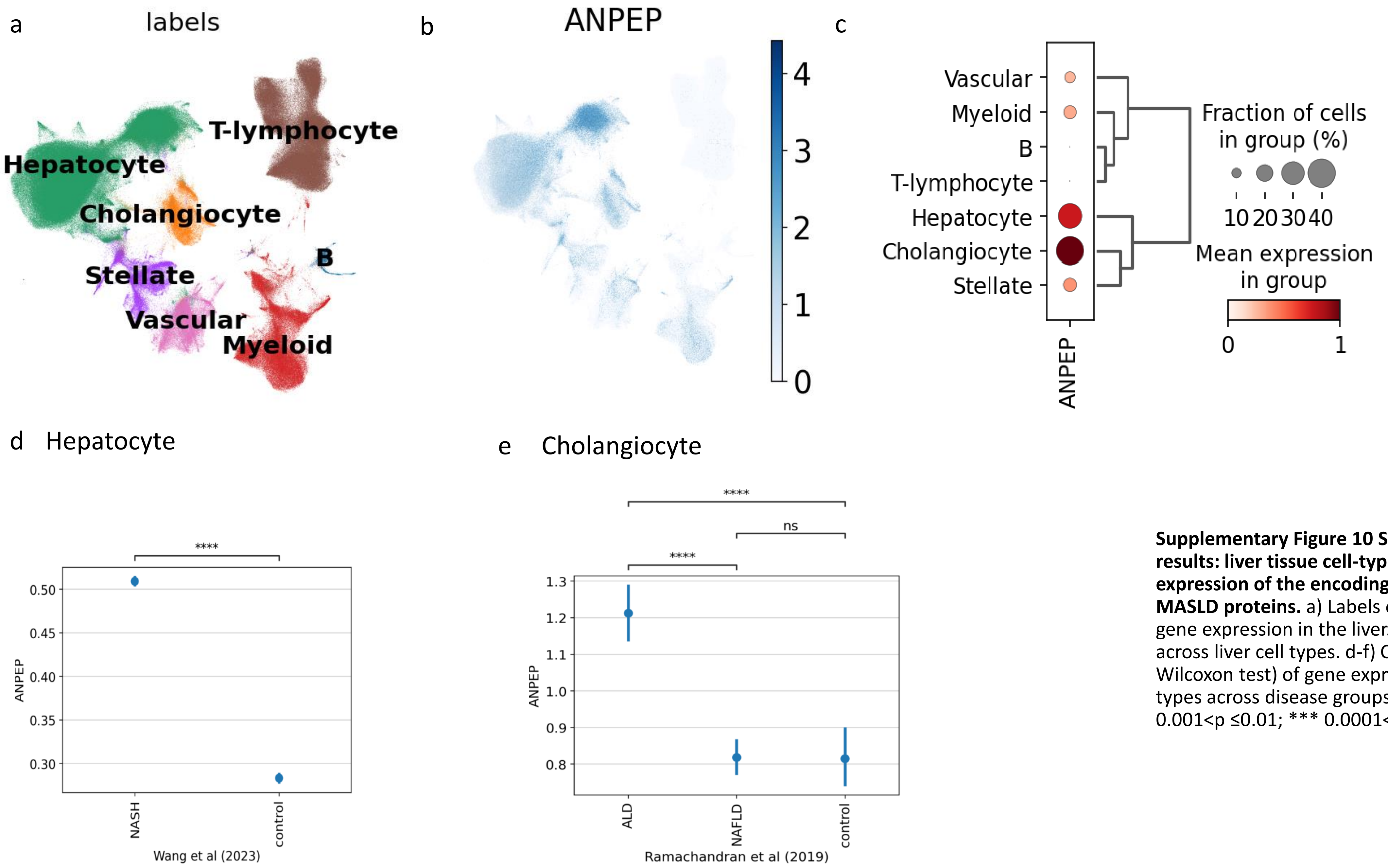


**Supplementary Figure 10 ScRNA-seq and snRNA-seq results: liver tissue cell-type-based differential gene expression of the encoding genes of the 17 candidate MASLD proteins.** a) Labels of UMAP. b) UMAP of the gene expression in the liver. c) Gene expression levels across liver cell types. d-f) Comparison (Mann-Whitney-Wilcoxon test) of gene expression levels in single cell types across disease groups. Ns, p≤1; * 0.01<p ≤0.05; ** 0.001<p ≤0.01; *** 0.0001<p ≤0.001; **** p ≤0.0001.

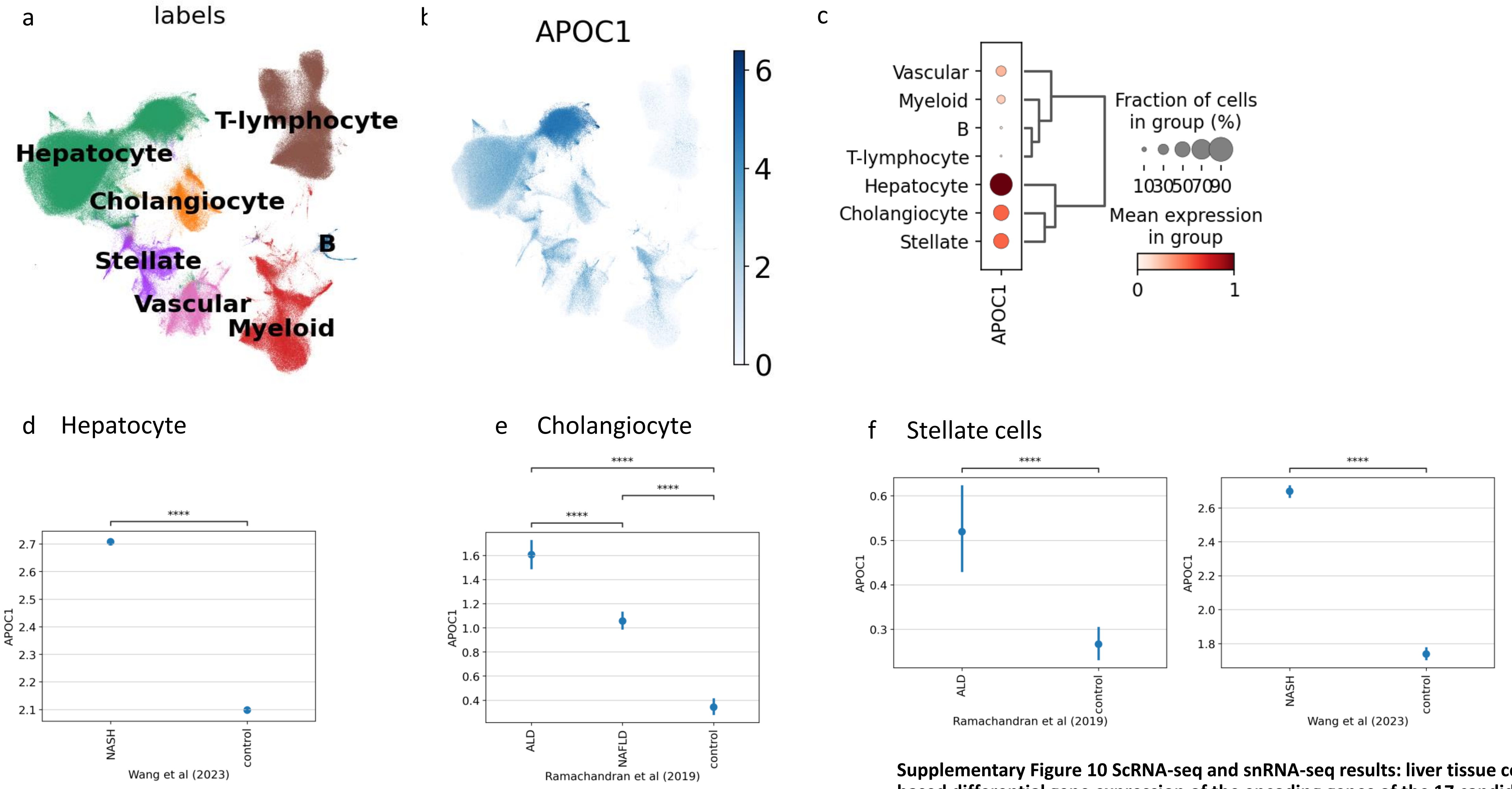


**Supplementary Figure 10 ScRNA-seq and snRNA-seq results: liver tissue cell-type-based differential gene expression of the encoding genes of the 17 candidate MASLD proteins.** a) Labels of UMAP. b) UMAP of the gene expression in the liver. c) Gene expression levels across liver cell types. d-e) Comparison (Mann-Whitney-Wilcoxon test) of gene expression levels in single cell types across disease groups. Ns, p≤1; * 0.01<p ≤0.05; ** 0.001<p ≤0.01; *** 0.0001<p ≤0.001; **** p ≤0.0001.

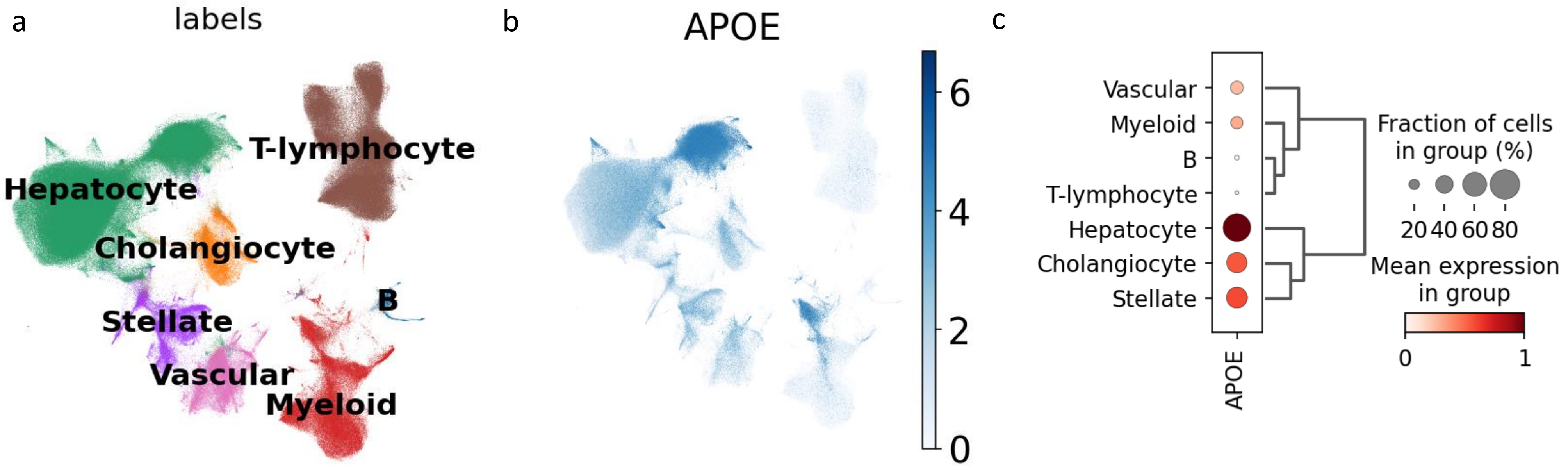


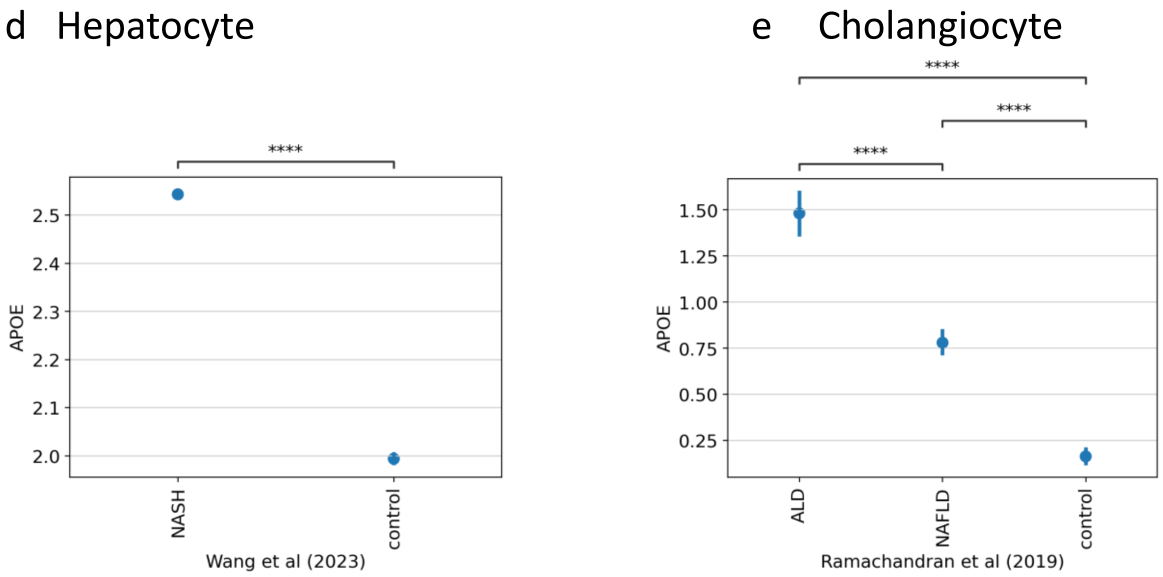


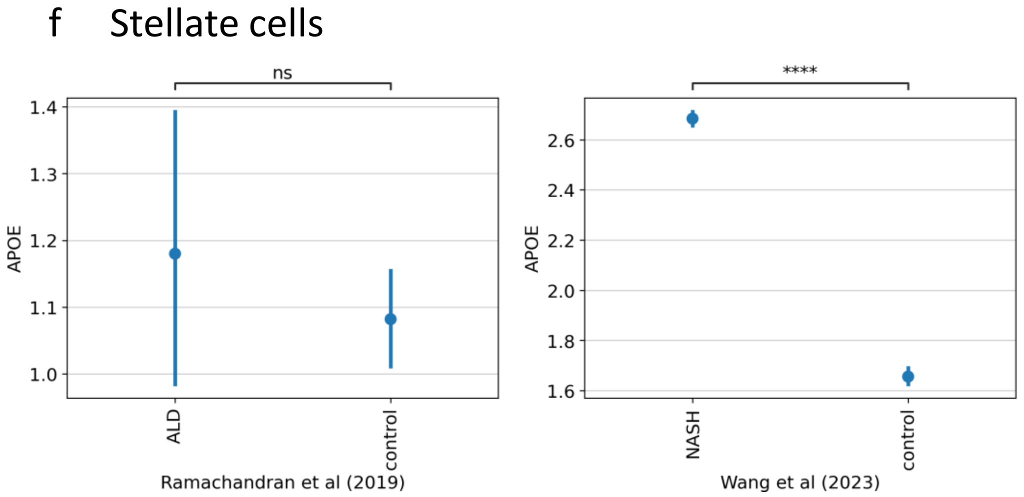


**Supplementary Figure 10 ScRNA-seq and snRNA-seq results: liver tissue cell-type-based differential gene expression of the encoding genes of the 17 candidate MASLD proteins.** a) Labels of UMAP. b) UMAP of the gene expression in the liver. c) Gene expression levels across liver cell types. d-e) Comparison (Mann-Whitney-Wilcoxon test) of gene expression levels in single cell types across disease groups. Ns, $p \le 1$; * $0.01 < p \le 0.05$; ** $0.001 < p \le 0.01$; *** $0.0001 < p \le 0.001$; **** $p \le 0.0001$.

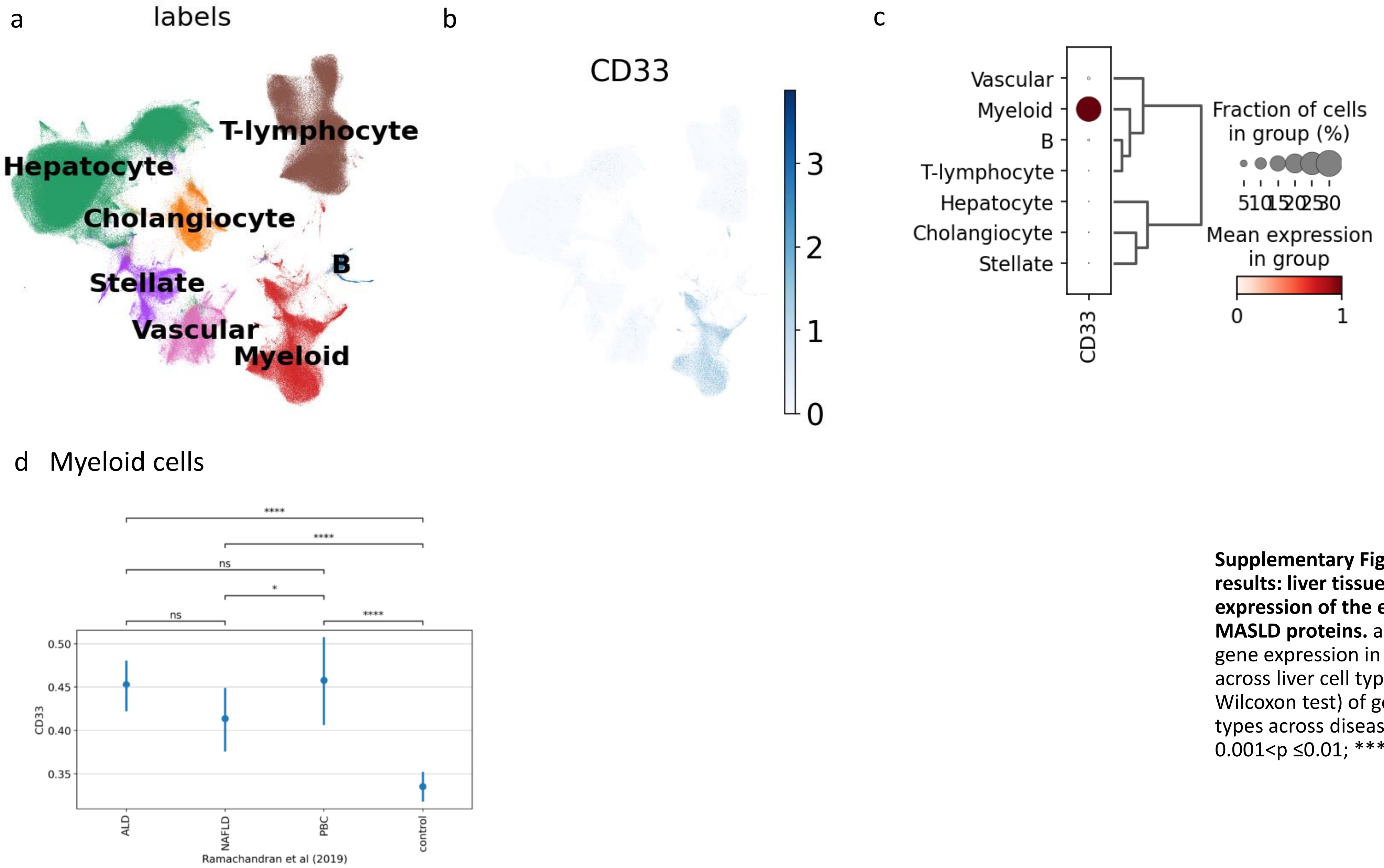


**Supplementary Figure 10 ScRNA-seq and snRNA-seq results: liver tissue cell-type-based differential gene expression of the encoding genes of the 17 candidate MASLD proteins.** a) Labels of UMAP. b) UMAP of the gene expression in the liver. c) Gene expression levels across liver cell types. d-e) Comparison (Mann-Whitney-Wilcoxon test) of gene expression levels in single cell types across disease groups. Ns, p≤1; * 0.01<p ≤0.05; ** 0.001<p ≤0.01; *** 0.0001<p ≤0.001; **** p ≤0.0001.

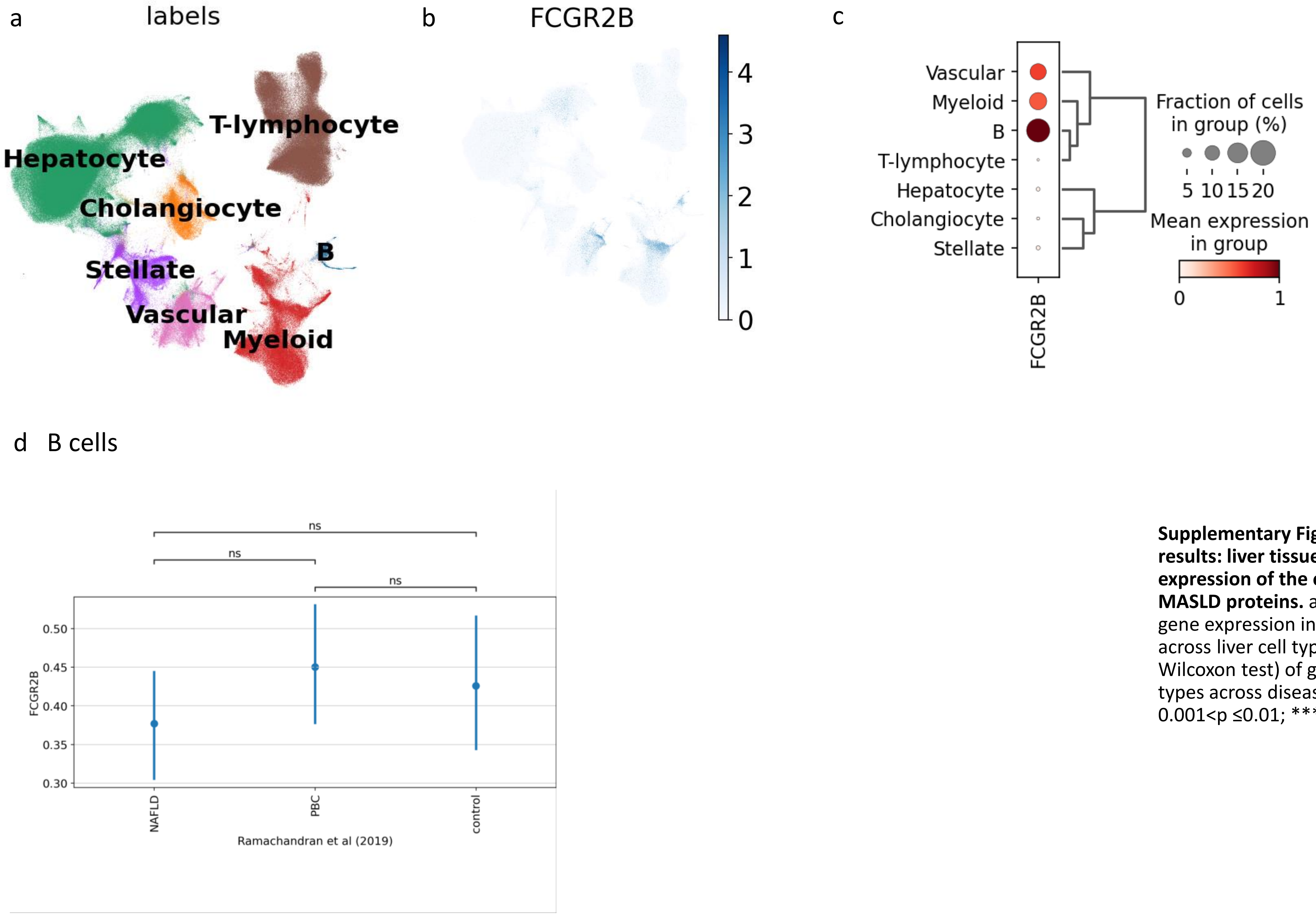


**Supplementary Figure 10 ScRNA-seq and snRNA-seq results: liver tissue cell-type-based differential gene expression of the encoding genes of the 17 candidate MASLD proteins.** a) Labels of UMAP. b) UMAP of the gene expression in the liver. c) Gene expression levels across liver cell types. d-e) Comparison (Mann-Whitney-Wilcoxon test) of gene expression levels in single cell types across disease groups. Ns, p≤1; * 0.01<p ≤0.05; ** 0.001<p ≤0.01; *** 0.0001<p ≤0.001; **** p ≤0.0001.

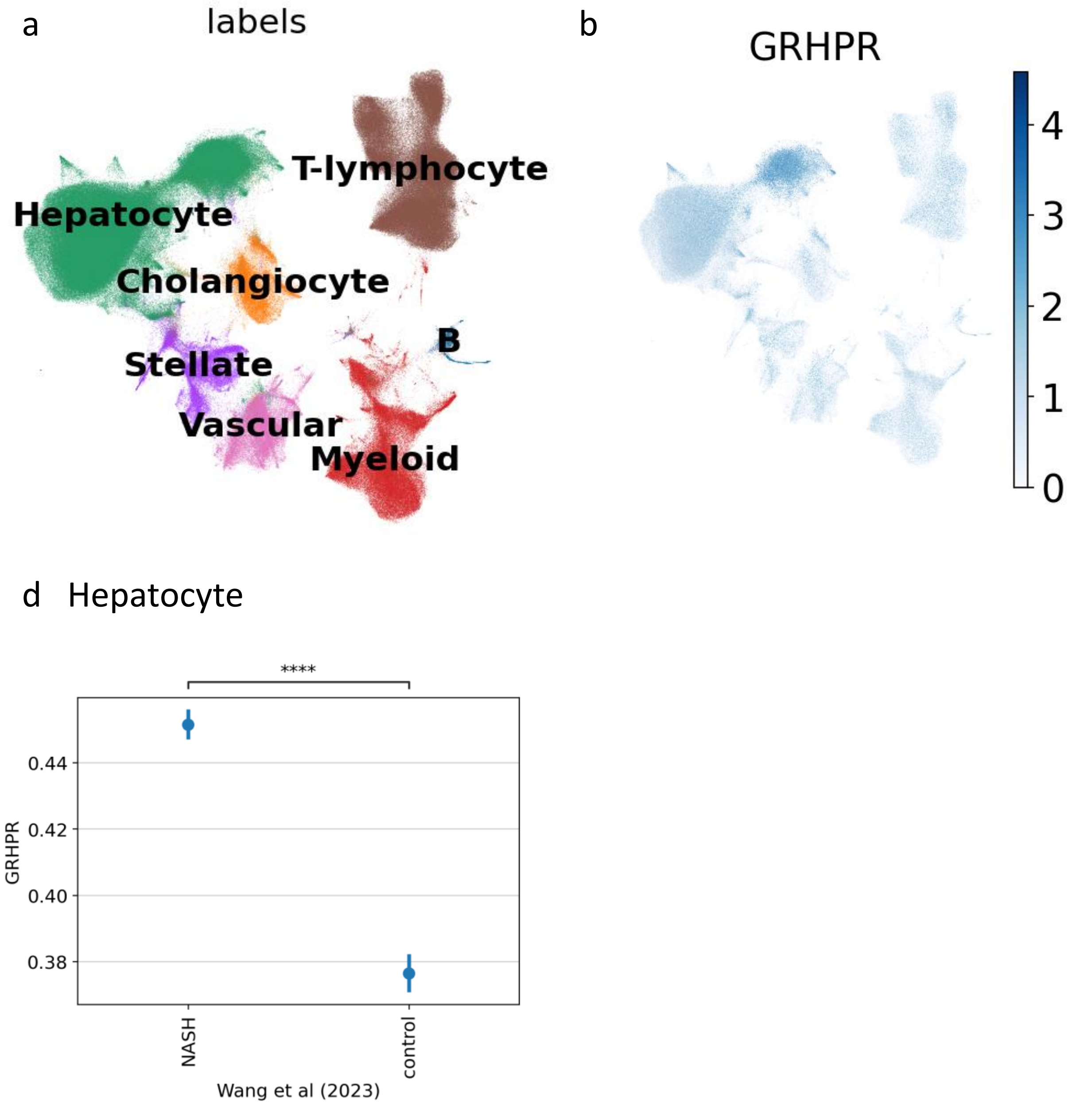


**Supplementary Figure 10 ScRNA-seq and snRNA-seq results: liver tissue cell-type-based differential gene expression of the encoding genes of the 17 candidate MASLD proteins.** a) Labels of UMAP. b) UMAP of the gene expression in the liver. c) Gene expression levels across liver cell types. d-e) Comparison (Mann-Whitney-Wilcoxon test) of gene expression levels in single cell types across disease groups. Ns, $p \leq 1$; * $0.01 < p \leq 0.05$; ** $0.001 < p \leq 0.01$; *** $0.0001 < p \leq 0.001$; **** $p \leq 0.0001$.

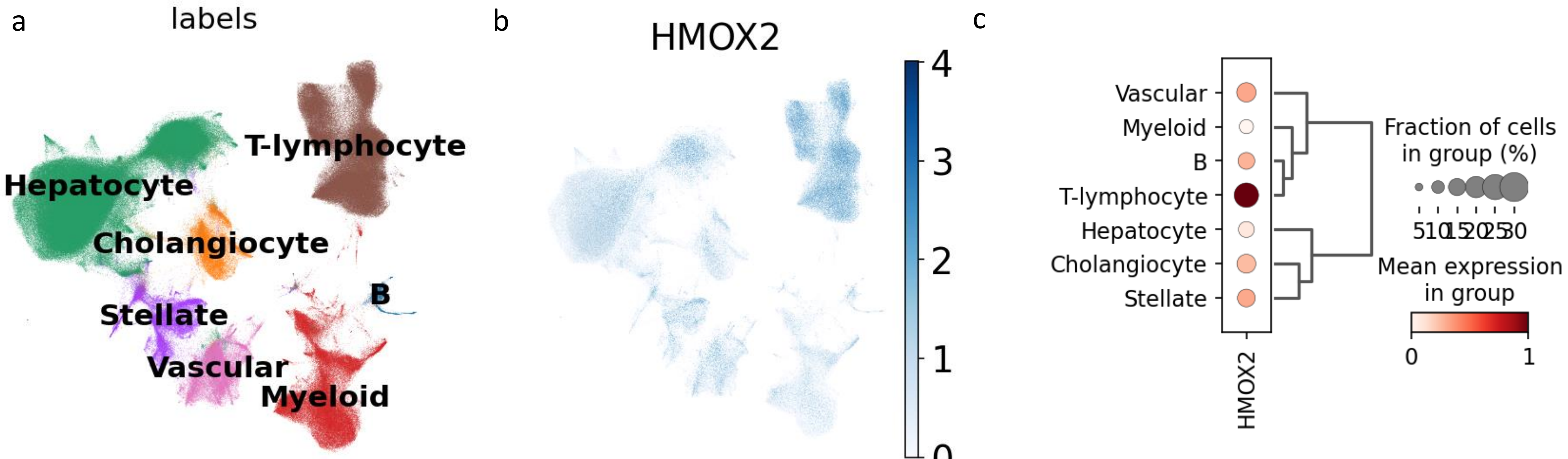


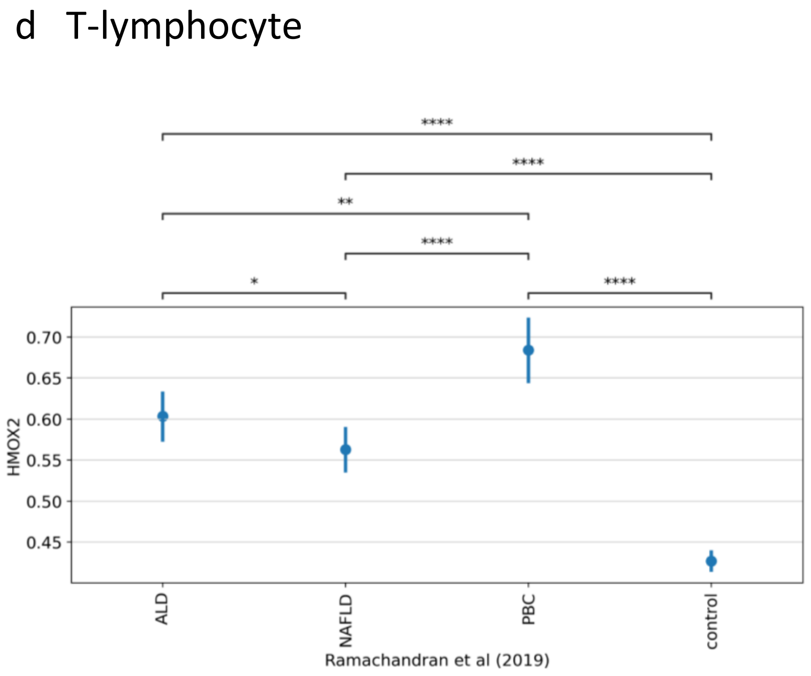


**Supplementary Figure 10 ScRNA-seq and snRNA-seq results: liver tissue cell-type-based differential gene expression of the encoding genes of the 17 candidate MASLD proteins.** a) Labels of UMAP. b) UMAP of the gene expression in the liver. c) Gene expression levels across liver cell types. d-e) Comparison (Mann-Whitney-Wilcoxon test) of gene expression levels in single cell types across disease groups. Ns, p≤1; * 0.01<p ≤0.05; ** 0.001<p ≤0.01; *** 0.0001<p ≤0.001; **** p ≤0.0001.

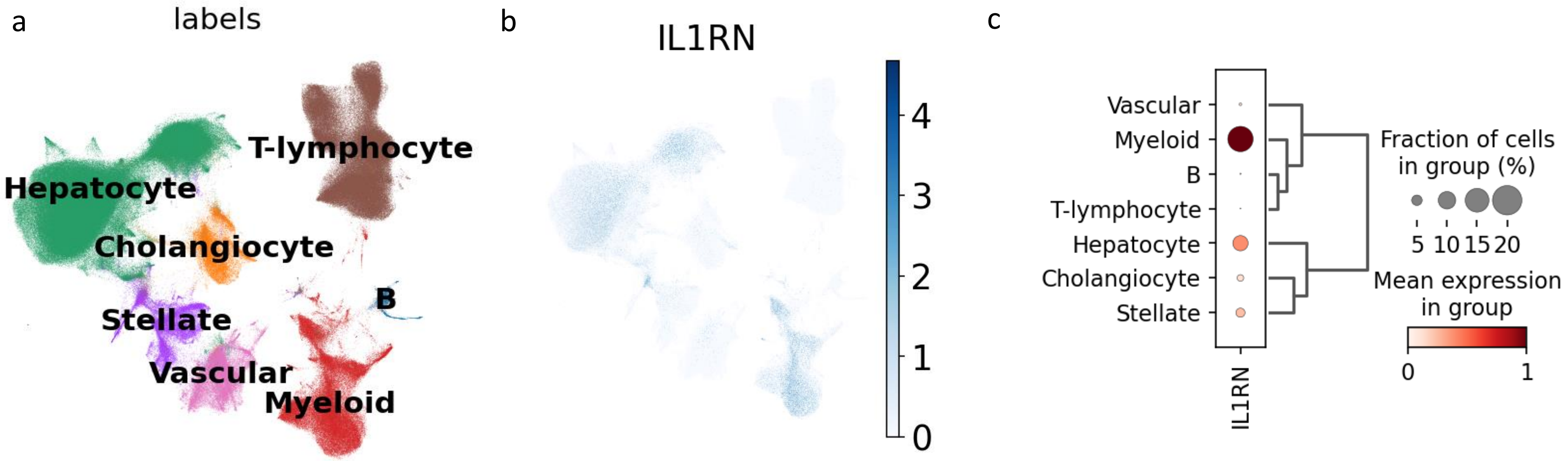


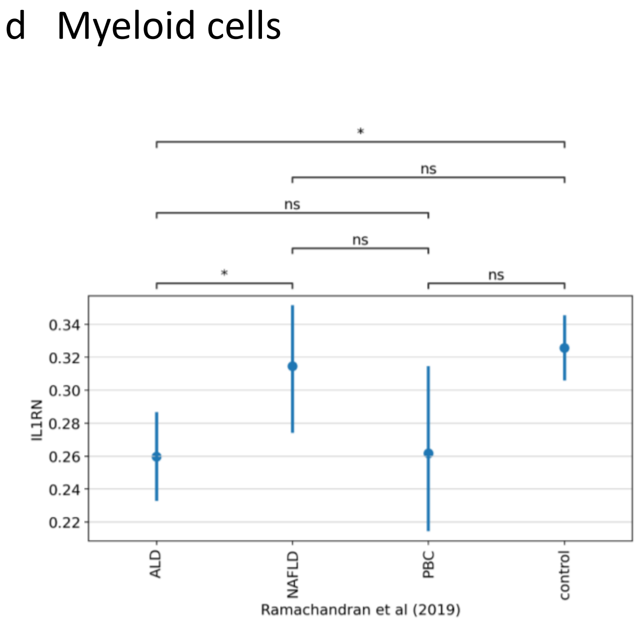


**Supplementary Figure 10 ScRNA-seq and snRNA-seq results: liver tissue cell-type-based differential gene expression of the encoding genes of the 17 candidate MASLD proteins.** a) Labels of UMAP. b) UMAP of the gene expression in the liver. c) Gene expression levels across liver cell types. d-e) Comparison (Mann-Whitney-Wilcoxon test) of gene expression levels in single cell types across disease groups. Ns, p≤1; * 0.01<p ≤0.05; ** 0.001<p ≤0.01; *** 0.0001<p ≤0.001; **** p ≤0.0001.

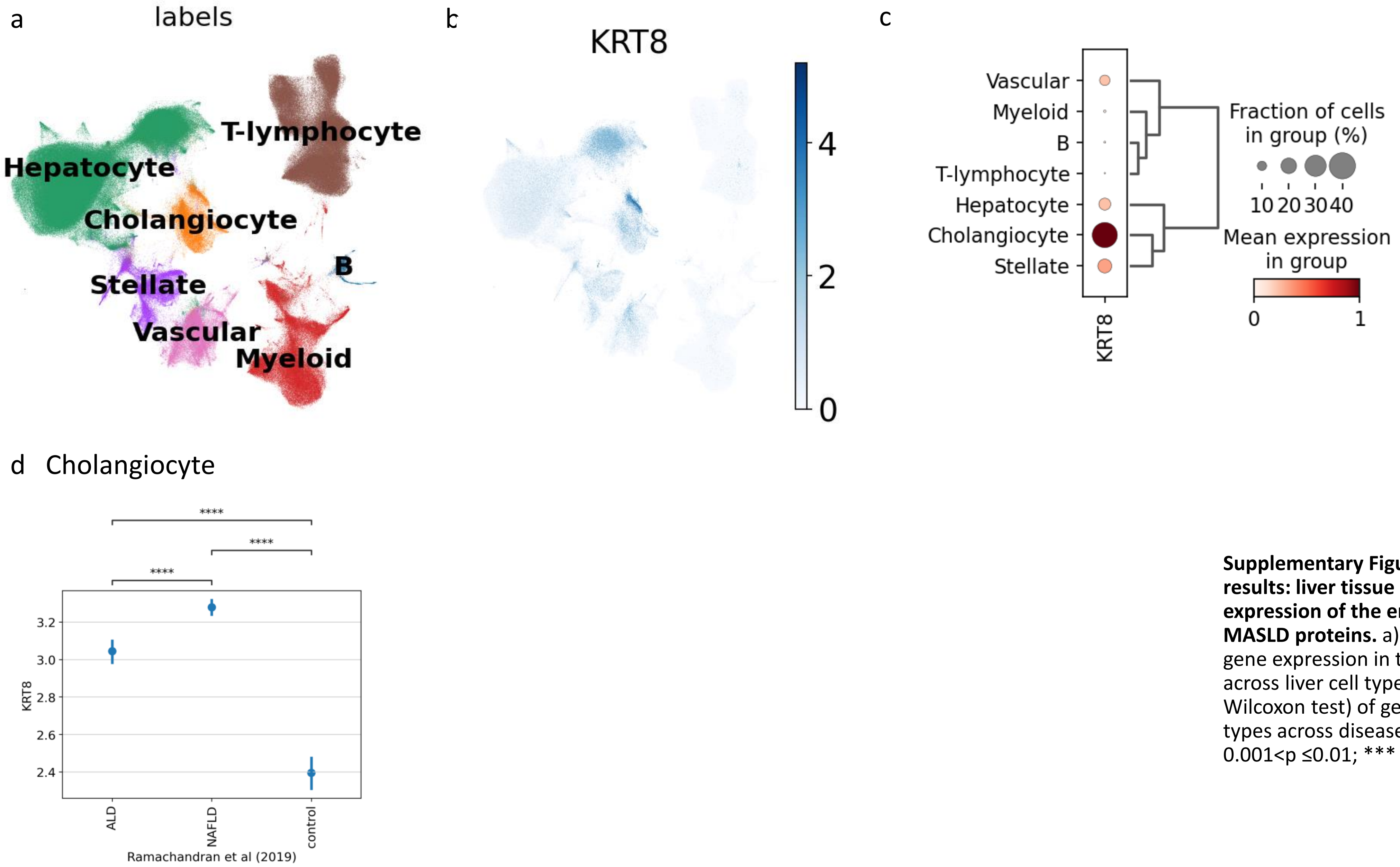


**Supplementary Figure 10 ScRNA-seq and snRNA-seq results: liver tissue cell-type-based differential gene expression of the encoding genes of the 17 candidate MASLD proteins.** a) Labels of UMAP. b) UMAP of the gene expression in the liver. c) Gene expression levels across liver cell types. d-e) Comparison (Mann-Whitney-Wilcoxon test) of gene expression levels in single cell types across disease groups. Ns, p≤1; * 0.01<p ≤0.05; ** 0.001<p ≤0.01; *** 0.0001<p ≤0.001; **** p ≤0.0001.

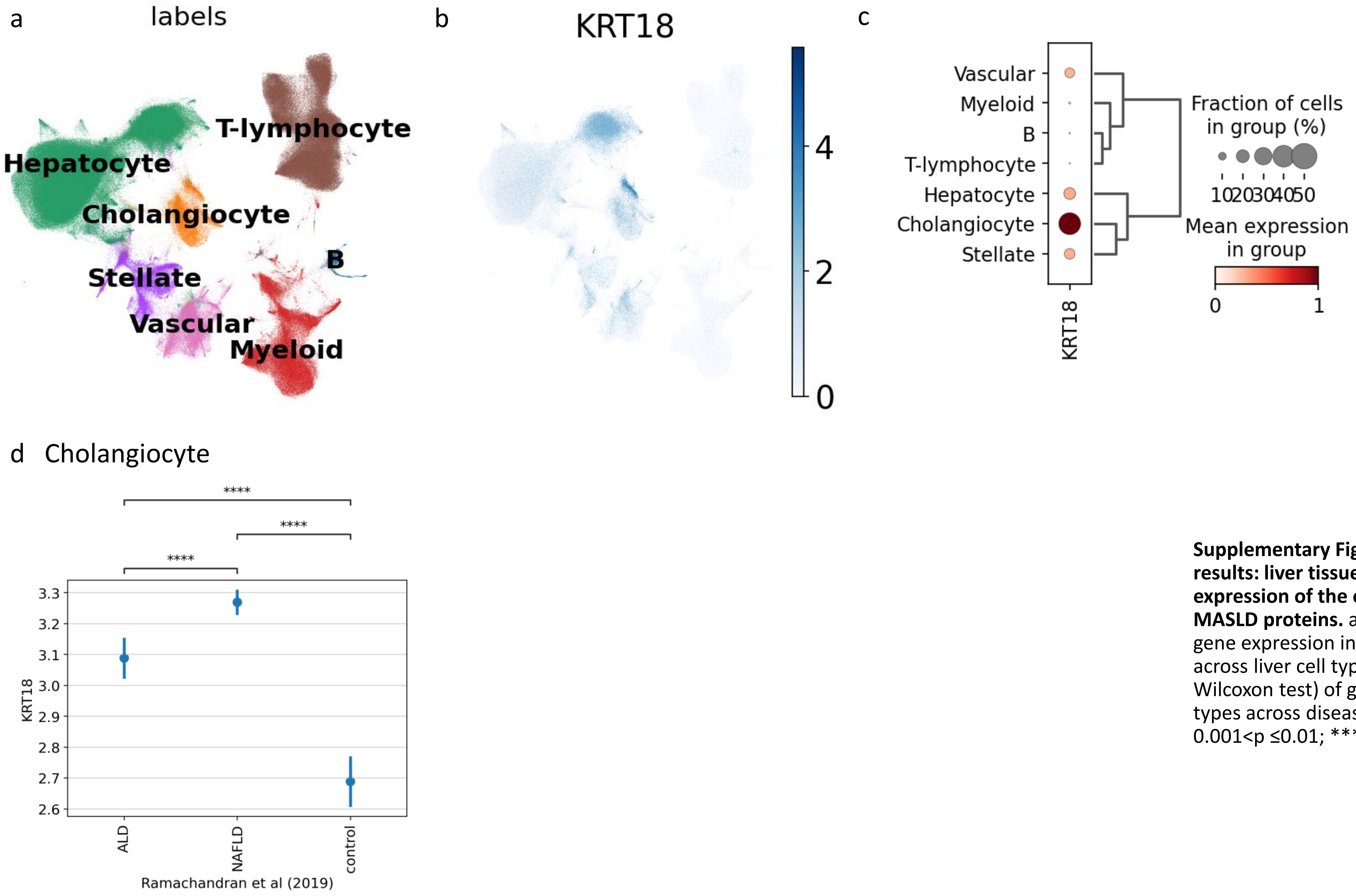


**Supplementary Figure 10 ScRNA-seq and snRNA-seq results: liver tissue cell-type-based differential gene expression of the encoding genes of the 17 candidate MASLD proteins.** a) Labels of UMAP. b) UMAP of the gene expression in the liver. c) Gene expression levels across liver cell types. d-e) Comparison (Mann-Whitney-Wilcoxon test) of gene expression levels in single cell types across disease groups. Ns, p≤1; * 0.01<p ≤0.05; ** 0.001<p ≤0.01; *** 0.0001<p ≤0.001; **** p ≤0.0001.

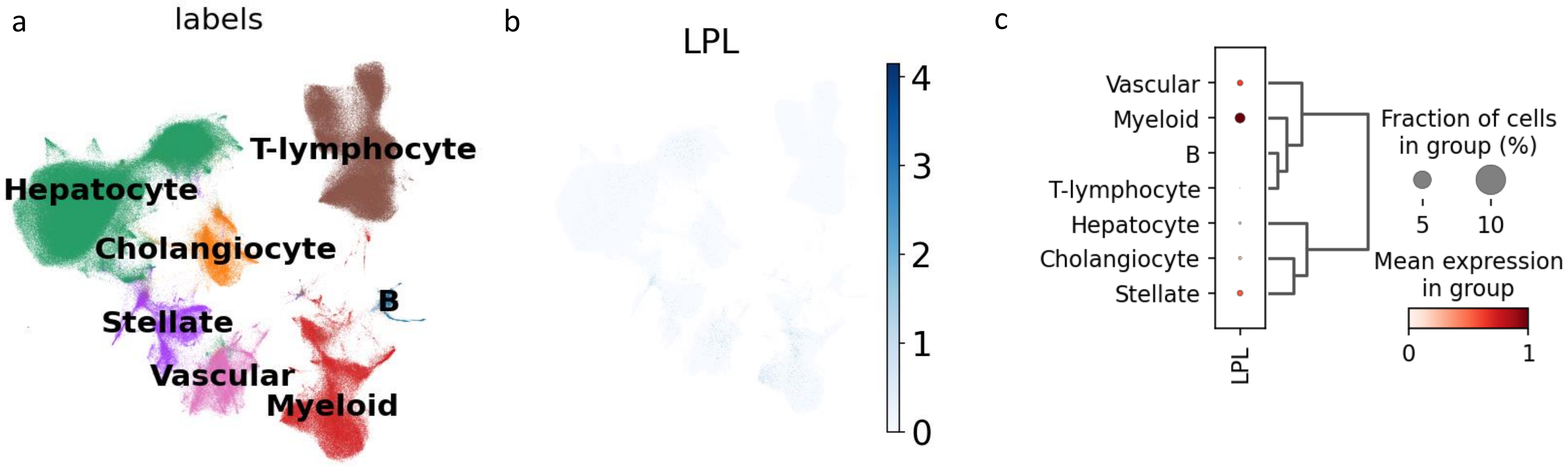


**Supplementary Figure 10 ScRNA-seq and snRNA-seq results: liver tissue cell-type-based differential gene expression of the encoding genes of the 17 candidate MASLD proteins.** a) Labels of UMAP. b) UMAP of the gene expression in the liver. c) Gene expression levels across liver cell types. d-e) Comparison (Mann-Whitney-Wilcoxon test) of gene expression levels in single cell types across disease groups. Ns, p≤1; * 0.01<p ≤0.05; ** 0.001<p ≤0.01; *** 0.0001<p ≤0.001; **** p ≤0.0001.

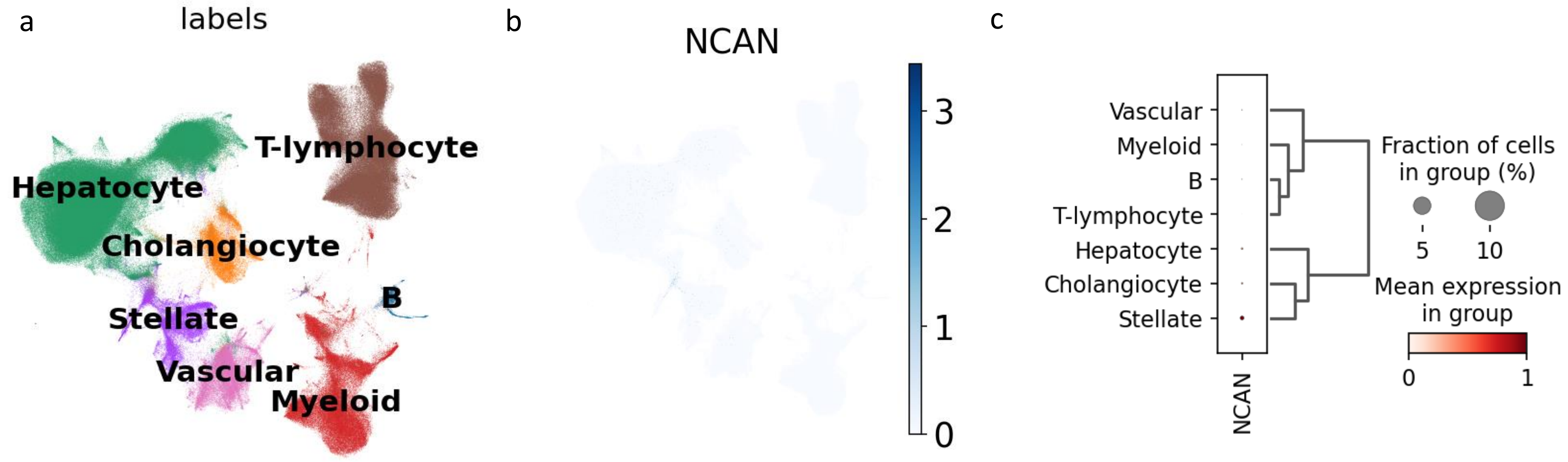


**Supplementary Figure 10 ScRNA-seq and snRNA-seq results: liver tissue cell-type-based differential gene expression of the encoding genes of the 17 candidate MASLD proteins.** a) Labels of UMAP. b) UMAP of the gene expression in the liver. c) Gene expression levels across liver cell types. d-e) Comparison (Mann-Whitney-Wilcoxon test) of gene expression levels in single cell types across disease groups. Ns, p≤1; * 0.01<p ≤0.05; ** 0.001<p ≤0.01; *** 0.0001<p ≤0.001; **** p ≤0.0001.

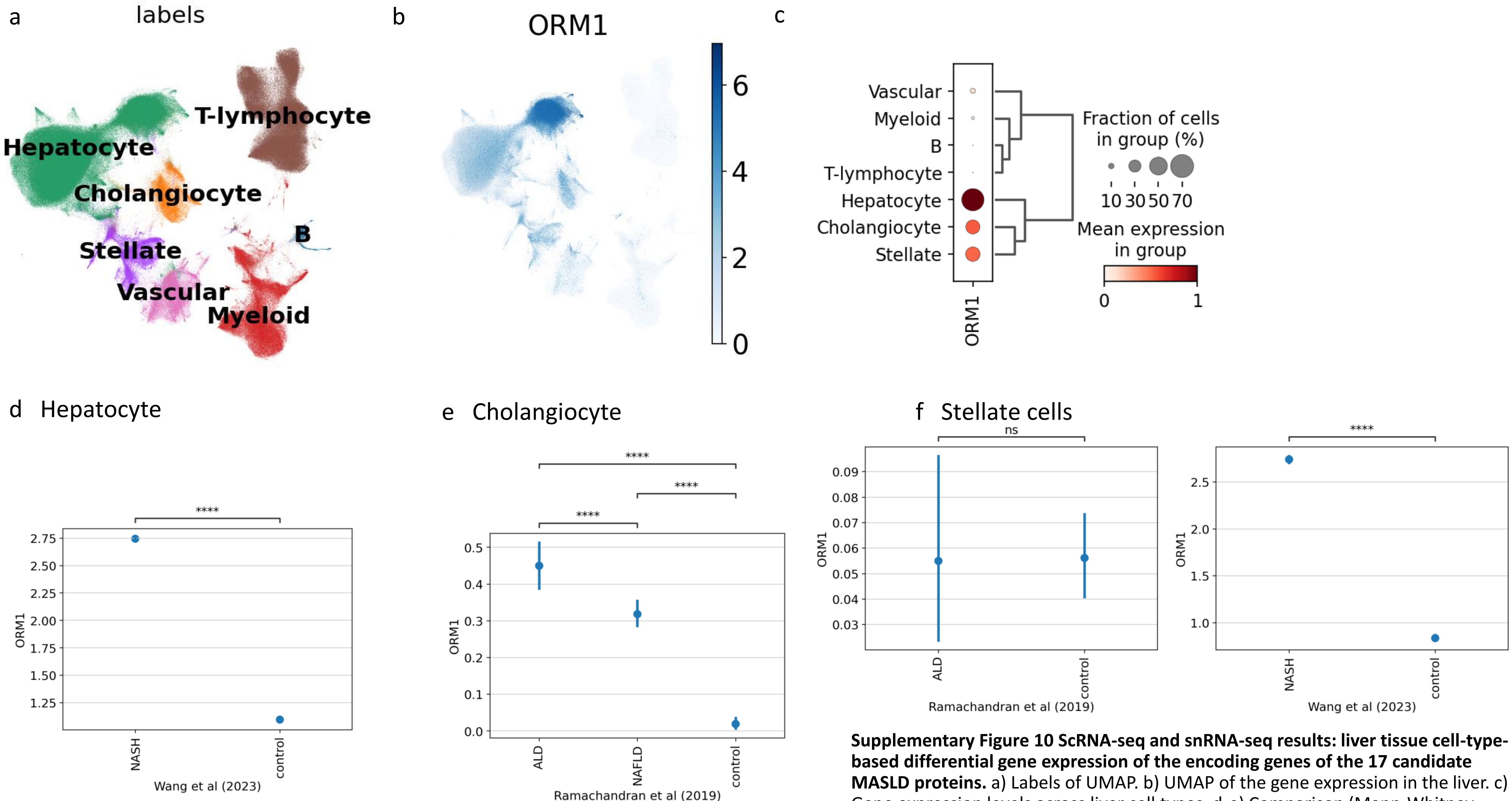


**Supplementary Figure 10 ScRNA-seq and snRNA-seq results: liver tissue cell-type-based differential gene expression of the encoding genes of the 17 candidate MASLD proteins.** a) Labels of UMAP. b) UMAP of the gene expression in the liver. c) Gene expression levels across liver cell types. d-e) Comparison (Mann-Whitney-Wilcoxon test) of gene expression levels in single cell types across disease groups. Ns, $p \leq 1$; * $0.01 < p \leq 0.05$; ** $0.001 < p \leq 0.01$; *** $0.0001 < p \leq 0.001$; **** $p \leq 0.0001$.

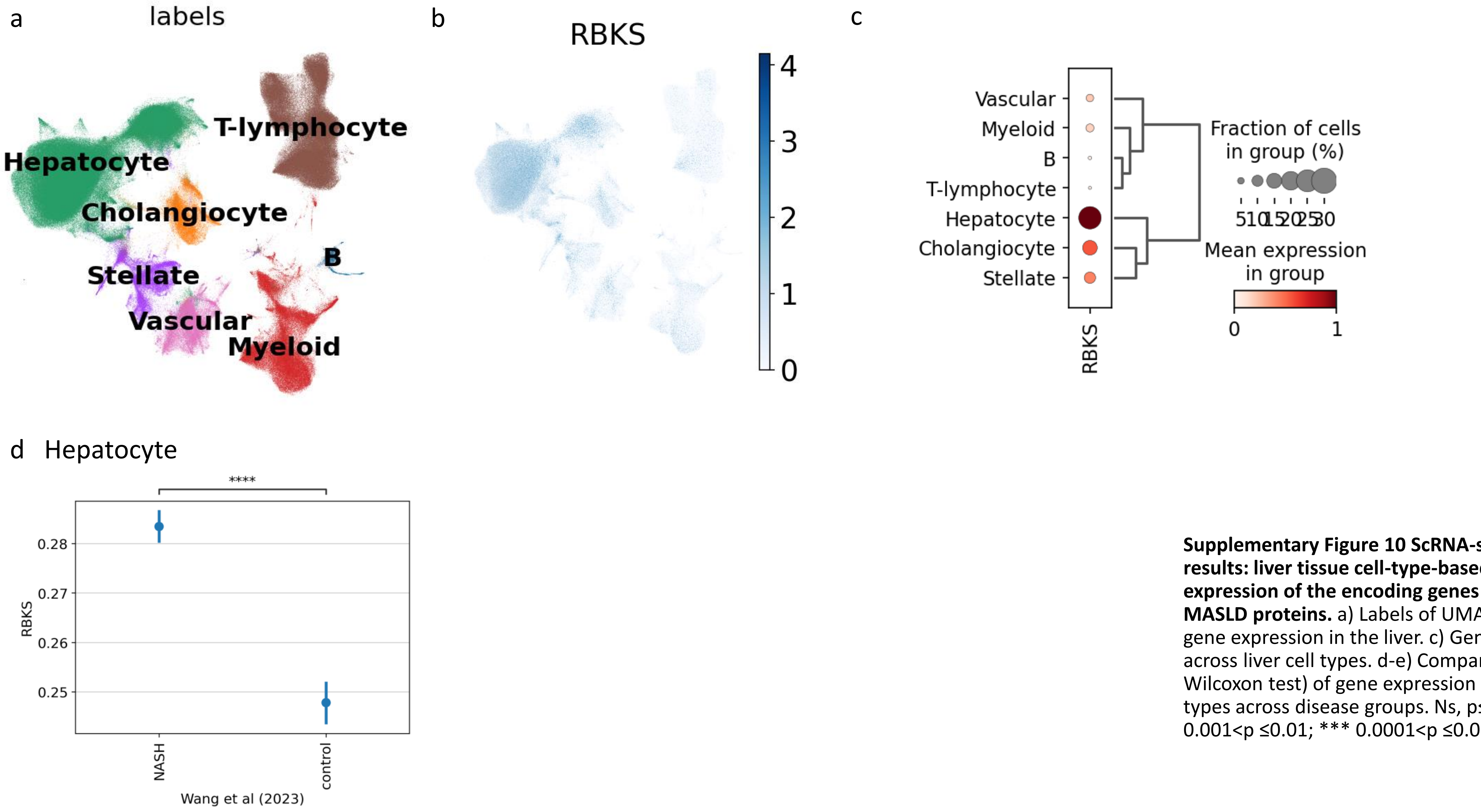


**Supplementary Figure 10 ScRNA-seq and snRNA-seq results: liver tissue cell-type-based differential gene expression of the encoding genes of the 17 candidate MASLD proteins.** a) Labels of UMAP. b) UMAP of the gene expression in the liver. c) Gene expression levels across liver cell types. d-e) Comparison (Mann-Whitney-Wilcoxon test) of gene expression levels in single cell types across disease groups. Ns, p≤1; * 0.01<p ≤0.05; ** 0.001<p ≤0.01; *** 0.0001<p ≤0.001; **** p ≤0.0001.

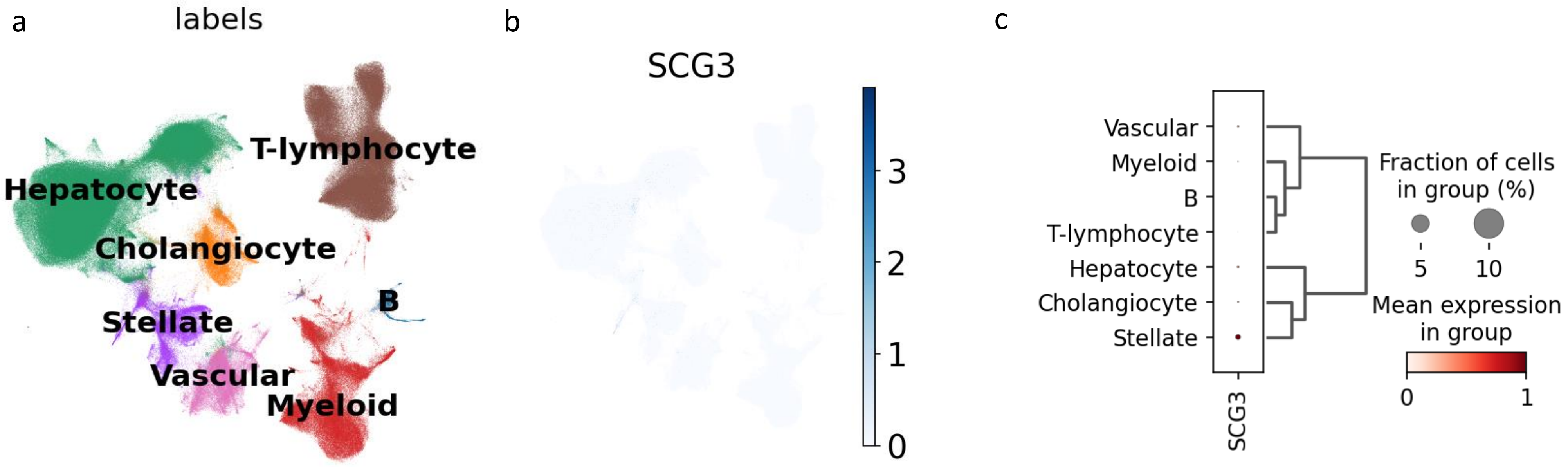


**Supplementary Figure 10 ScRNA-seq and snRNA-seq results: liver tissue cell-type-based differential gene expression of the encoding genes of the 17 candidate MASLD proteins.** a) Labels of UMAP. b) UMAP of the gene expression in the liver. c) Gene expression levels across liver cell types. d-e) Comparison (Mann-Whitney-Wilcoxon test) of gene expression levels in single cell types across disease groups. Ns, p≤1; * 0.01<p ≤0.05; ** 0.001<p ≤0.01; *** 0.0001<p ≤0.001; **** p ≤0.0001.

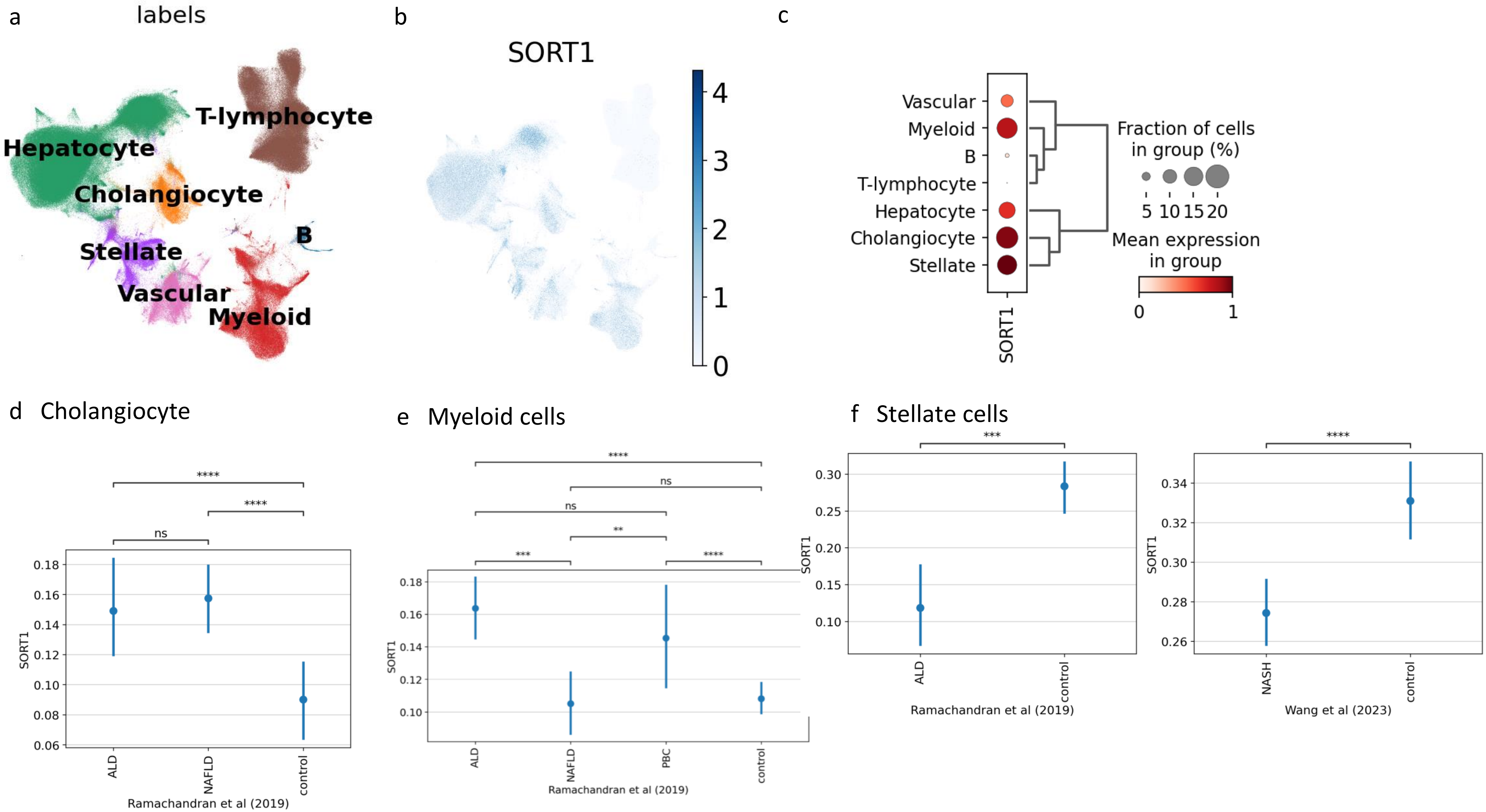
a
labels
T-lymphocyte
Hepatocyte
Cholangiocyte
B
Stellate
Vascular
Myeloid
b
SORT1
4
3
2
1
0
c
Vascular
Myeloid
B
T-lymphocyte
Hepatocyte
Cholangiocyte
Stellate
SORT1
Fraction of cells
in group (%)
5 10 15 20
Mean expression
in group
0
1
d Cholangiocyte
****
****
ns
SORT1
0.18
0.16
0.14
0.12
0.10
0.08
0.06
ALD
NAFLD
control
Ramachandran et al (2019)
e Myeloid cells
****
ns
ns
**
***
****
SORT1
0.18
0.16
0.14
0.12
0.10
ALD
NAFLD
PBC
control
Ramachandran et al (2019)
f Stellate cells
***
SORT1
0.30
0.25
0.20
0.15
0.10
ALD
control
Ramachandran et al (2019)
****
SORT1
0.34
0.32
0.30
0.28
0.26
NASH
control
Wang et al (2023)

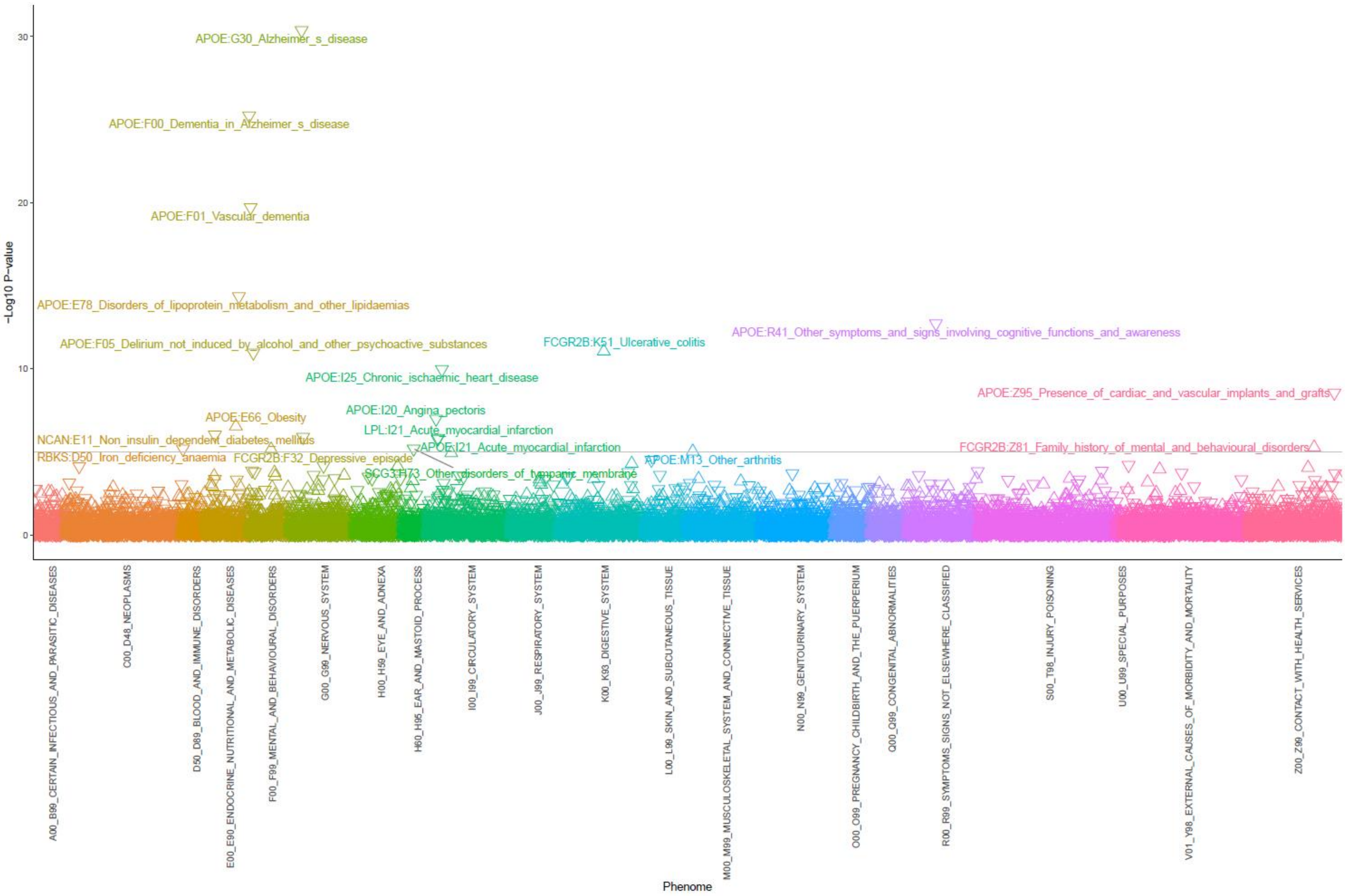


**Supplementary Figure 11 MR-PheWAS results of the 17 candidate MASLD proteins.** Proteins with 3 digits ICD-10 codes and disease names are shown for the significant associations ($p<1\times10^{-5}$). △ Positive association of protein and phenotype by MR. ▽ Negative association of protein and phenotype by MR.
