## Supplementary Materials for "Profiling the genome and proteome of metabolic dysfunction-associated steatotic liver disease identifies potential therapeutic targets"

**Proteomics analysis of liver biopsies**

*Liver tissue homogenisation and protein digestion*

All procedures were performed with pre-cooled reagents, tubes and instruments at 4 °C. Approximately 100 mg wet tissue weight of liver tissue was removed from -80°C storage and placed in homogenising tube (Bertin Instruments) which was pre-filled with ~100 µl of 1.4mm ceramic (zirconium oxide) beads. Immediately, 700 µl of lysis buffer consisting of 8 M urea in 50 mM ammonium bicarbonate and protease inhibitor cocktail (0.1% v/v) (P8340, Sigma Aldrich) was added to the tissue. The samples were homogenised using a Precellys 24 homogeniser (Bertin Instruments) for two 20 second bursts at 5500 RPM, with a 30 second pause between bursts. The lysates were transferred to new Eppendorf Lobind tubes. Protein concentration was determined by bicinchoninic acid assay (Pierce). For protein digestion, 100 μg of each sample was aliquoted and volumes were normalized to 23 µl with additional lysis buffer. Each sample was reduced, alkylated, cleaned down and digested on column with S-Trap^TM^ mini MS sample kit: 100-300ug (Protifi, USA) according to the manufacturer’s instructions. Following digestion, peptides were eluted stepwise with 3 elution buffers including 50 mM TEAB in water, 0.2% formic acid in water, and 50% acetonitrile/0.2% formic acid in water at a volume of 80 μL each. Eluted peptides were dried on an evaporating centrifuge and reconstituted to 1 µg/µl in 0.1% formic acid, vortexed and sonicated for 10 minutes before centrifugation at 10,000 x g for 5 minutes. 8 µl of the supernatant was transferred to an HPLC vial for individual sample pre-fractionation.

Pre-fractionation was carried out using a Thermo Ultimate 3000 high performance liquid chromatography (HPLC), equipped with an inline degasser, autosampler with integrated fraction collector, column oven, UV/visible wavelength diode array detector. An ACQUITY UPLC CSH 1.7 µm 1.0 x 150 mm column (Waters, USA) was used for separation, with the column temperature maintained at 35°C. The flow rate was 30 µl/min. The gradient started at 95% solvent A and 5% solvent B, and was held constant for 5 minutes. The concentration of solvent B increased to 50% over 32 minutes. Peptides were eluted over the 50 minutes period and 10 separate fractions were collected between 4 and 44 minutes. Spectra were collected at 214 nm to determine peptide elution times. These fractions were pooled into five fractions using a concatenation strategy, whereby every fifth fraction was pooled. The five fractions were dried and resuspended in 6 µl 0.1% formic acid in water.

*DDA-MS data acquisition*

The fractions were analysed by liquid chromatography-tandem mass spectrometry (LC-MS/MS) in DDA mode. All mass spectrometry runs were performed on a Thermo Scientific Easy-nLC 1200 high performance liquid chromatography (HPLC), coupled to a Thermo Scientific Q-Exactive HF-X mass spectrometer. Samples (1 µg in 5 µl) were injected onto a reversed-phase capillary column (Thermo Scientific PepMap RLSC C18 2 µm, 100A, 75µm x 50 cm) that was maintained at 50°C. The flow rate was 0.3 µl/min. The gradient of solvent B (80% acetonitrile in water with 0.1% formic acid) started from 5% and increased to 35% over 85 min, to 55% over 21 minutes, to 80% over 2 minutes, held at 80% for 2 minutes, followed by a 6 minute column equilibration step at 95% A (0.1% formic acid in water). The liquid chromatography eluent was analysed using a Thermo Q-Exactive HF-X system equipped with an EasySpray source and controlled with Xcalibur 4.2.47 software.

Peptide spectra were acquired with a Top 20 data-dependent MS/MS scan method; the parameters were set as follows: Q-Exactive HF-X operated in positive ionization mode with default charge set as 2+, charge states more than 8+ and less than 1+ were excluded from the analysis. The range of full-MS scan was 350–1400 m/z with resolving power 30,000 at m/z 200 and acquisition gain control target set as 3E6 with 54 ms maximum injection time. Tandem mass spectra were acquired with scan range starting from 100 first fixed mass and dynamically calculated maximum m/z dependent on the precursor ion m/z. The MS/MS resolving power was set as 60,000 at m/z 200 and acquisition gain control target was determined as 5E5 with 118 ms maximum injection time. The precursor ion was isolated to MS/MS scan in narrow isolation window of 1.3 m/z with 0.0 m/z offset. Precursor ions were dynamically excluded for 20 s after MS/MS scan with 2 ppm precision. The maximum number of precursor ions allowed to be fragmented in one duty cycle was 20 (loop count 20) with normalized collision energy of 28.

*LC-MS Data analysis*

Raw MS data were processed with PEAKS Xpro Studio Software 10.6. Protein sequences of the complete human proteome provided by Uniprot (June 2021, 202152 sequences) was used for protein identification. MS data files were subjected to default data refinement. The parent mass error tolerance was set to 10 and the fragment mass error tolerance to 0.02 Da. Enzyme was specified as trypsin, and a maximum of 3 missed cleavages allowed. Methyl methanethiosulfonate dithiomethane was set as a fixed modification on cysteine residues. Oxidation of methionine and acetylation of lysine residues were set in the de novo and database peptide searches as variable PTMs. A 1% FDR cutoff was applied, and all peptides identified by PEAKS DB were defined as linear peptides. Further data analysis was performed in R Studio Workbench 1.4.1717-3.

**Data preparation**

Data preparation of continuous variables was based on the overall population in UK Biobank. They were first log-transform if skewed by eye or skewness value out of 2. The extreme values were replaced with the minimum or maximum (mean+/-5×standard deviation) values.

To reduce subjects’ missingness due to missing partial variables, we performed a fast imputation of missing values by chained random forests through the R package *missRanger* to impute missing values for covariates, all of which had less than 20% missing. Information used in the imputation model included age, sex, fasting time, smoking status, pack-years, alcohol frequency, alcohol grams per week, education, physical activity, type 2 diabetes, AST, ALT, GGT, BMI, waist circumference, systolic blood pressure, diastolic blood pressure, anti-hypertensives, triglycerides, HDL, lipid-lowering medications, glucose, HbA1c, anti-diabetics, CRP, platelet, and albumin. Briefly, the large matrix was imputed with a maximum of ten chaining interactions and 200 trees and weighted by the number of non-missing values; three candidate non-missing values were selected from the predictive mean matching steps. We did not impute protein values and directly measured outcomes (i.e., MASLD) in our study.

**Significance levels for multiple testing**

Considering the correlation of the proteins, we defined the significance levels based on the Matrix Spectral Decomposition (MSD) method^1^ in which we estimated the number of independent vectors using the pairwise bivariate correlation matrix of the tests we performed. We defined the significance threshold of *p*<3.35×10^-5^ for proteome-wide association studies, which was corrected for the 1,491 independent vectors calculated from the 2,941 tested proteins (0.05/1,491); we defined the significance threshold of *p*<1.2×10^-4^ for analyses of the 824 MASLD proteins with MR available correcting for the 417 independent vectors (0.05/417); we defined the significance threshold of *p*<3.3×10^-3^ for analyses of the 17 candidate MASLD proteins correcting for the 15 independent vectors (0.05/15); we defined the significance threshold of *p*<8.3×10^-3^ for analyses of 6 MASLD proteins available in Samalogic platform and 8 MASLD proteins available in liver biopsies analsyis correcting for the 6 independent vectors (0.05/6).

**Analysis of snRNA-seq and scRNA-seq**

Count matrices from GEO accessions GSE136103, GSE185477, GSE189175, GSE189600, GSE192740, and GSE212837 were concatenated preserving common gene across cohorts^2–8^. We filtered out barcodes at the 1% sequencing depth extreme and with more than 25% of mitochondrial transcripts. We then identified the liver comprising cell types by clustering a cell nearest neighbour graph using the Leiden method. This cell graph was build based on cell-to-cell similarities in a PCA latent representation where technical effects had been accounted. PCA cell embedding was based on the top 1000 most variable genes across all cells. Cells were embedded in the top 30 principal components. We used the Harmony method^9^ to remove library specific effects in this 30PCs representation. Cell embedding and clustering were done using scanpy v1.9.3^10^. Cell type identity was defined by annotating the least granular clustering resolution with the highest average silhouette score. Cell cluster specific genes were identified using the Mann-Whitney-Wilcoxon test and were contrasted with well-known cell-type specific genes to define the cluster cell identity.
